# Suicidality phenotypes reflect both shared and distinct genetic factors

**DOI:** 10.64898/2026.05.14.26353207

**Authors:** Sarah MC Colbert, Shane O’Connell, Howard J Edenberg, Nina Fajs, Emma C Johnson, Séverine Lannoy, Sandra Sanchez-Roige, Silviu-Alin Bacanu, Zuriel Ceja, Alexis C Edwards, Melanie E Garrett, Seonggyun Han, Eric T Monson, Emily K Roberts, Vladimir Vladimirov, Cynthia M Bulik, Brenda Cabrera-Mendoza, Christal N Davis, Giuseppe Fanelli, Ian C Fischer, Henry Fox-Jurkowitz, Gabriel R Fries, Marie E Gaine, Jose Guzman-Parra, Maria Koromina, Stefan Kloiber, Henry R Kranzler, Divya Mehta, John I Nurnberger, Mallory Stephenson, Fabian Streit, Claudio Toma, Alja Videtic Paska, Suicide Working Group of the Psychiatric Genomics Consortium, Nathan A Kimbrel, Allison E Ashley-Koch, Douglas Ruderfer, Anna R Docherty, Alexander S Hatoum, Niamh Mullins

## Abstract

Suicidality phenotypes, including suicidal ideation (SI), non-fatal suicide attempt (SA), and suicide death (SD), are heritable and exhibit both shared and phenotype-specific genetic influences. Using genomic structural equation modelling, we estimated the shared genetic architecture across GWAS of SI (176,147 cases, 1,010,300 controls), SA (53,919 cases, 1,063,988 controls), and SD (7,584 cases, 652,070 controls) and conducted a multivariate GWAS of a latent suicidality factor capturing their shared liability. This analysis identified 36 genome-wide significant loci, including seven not previously reported in any suicidality GWAS. Follow-up analyses identified residual genetic variance specific to each phenotype, including three SD-specific genomic risk loci. Conditioning suicidality phenotypes on genetic liability to psychiatric disorders revealed significant residual genetic variance across SI, SA, SD, and the suicidality common factor. Together, these results suggest that suicidality reflects both shared genetic liability and phenotype-specific contributions.

## MAIN

Suicidality is a major public health concern and a leading cause of death worldwide^1–5^. Suicidality includes a range of related but distinct phenotypes, including suicidal ideation (SI), non-fatal suicide attempt (SA), and suicide death (SD), which differ in prevalence, severity, and clinical presentation. Although these phenotypes are often conceptualized as reflecting increasing severity along a continuum, transitions between them are not strictly linear, with both overlapping and distinct risk factors characterizing these phenotypes. Consistent with ideation-to-action theories^6–9^, the emergence of SI and the progression to SA or SD are thought to involve distinct risk factors and processes, such as acquired capability to act on suicidal thoughts. Furthermore, twin and family studies indicate that suicidality is moderately heritable (30-55%), with evidence for both shared genetic liability across SI, SA, and SD, and phenotype-specific genetic influences underlying their differentiation^10–13^. Related behaviors, such as non-suicidal self-injury (NSSI), have been proposed to contribute to acquired capability, although evidence is mixed as to whether NSSI is genetically distinguishable from suicidality phenotypes^14–17^. Elucidating shared and distinct etiological factors that contribute to suicidality phenotypes is critical for understanding why some individuals transition from SI to SA or even SD and for improving risk stratification, prevention, and intervention.

Recent large-scale genome-wide association study (GWAS) meta-analyses of SI, SA, and SD from the Psychiatric Genomics Consortium Suicide Working Group (PGC SUI) have begun to characterize the genetic architectures of suicidality phenotypes^18^. These GWAS identified significant SNP-based heritability for SI, SA, and SD (*h^2^_SNP_* = 2.0-6.7%), both shared and phenotype-specific loci, and strong but incomplete genetic correlations across these suicidality phenotypes (r_g_ = 0.70-0.88)^18^. Suicidality phenotypes also show significant positive genetic correlations with psychiatric disorders^18^, such as depression^19–22^, post-traumatic stress disorder (PTSD)^23^, and substance use disorders^24^, although the magnitude of these associations varies, highlighting the complex and pleiotropic nature of their genetic architectures.

To date, most GWAS of suicidality have analyzed phenotypes either in isolation^18,23,25^ or in aggregated forms, such as suicidal behavior (SB; including SA and SD cases)^20,26^ or suicidal thoughts and behaviors (including SI, SA, and SD cases)^27^. While these approaches have yielded important insights, they do not fully capture the multivariate structure of suicidality, which is characterized by both shared liability and meaningful distinctions across phenotypes. For example, although SI often precedes SA, less than a third of individuals with SI attempt suicide^28^, suggesting partially distinct etiological pathways. This distinction is supported by epidemiological, family, and molecular genetic studies showing both overlapping and independent risk factors for SI and SA^29–33^. Similarly, SD represents a heterogeneous outcome that may involve additional or different contributions from genetic and environmental risk factors relative to SI and SA^13,34–37^. Therefore, univariate GWAS and traditional meta-analytic approaches are not well-suited to disentangle the overlapping and distinct genetic influences.

Genomic structural equation modelling (genomic SEM) provides a framework to address these limitations by modeling the shared and unique genetic architecture of related phenotypes using GWAS summary statistics^38^. By leveraging the genetic covariance matrix for a set of phenotypes, genomic SEM identifies latent factors that capture common genetic liability, while also estimating phenotype-specific residual genetic variance. A latent factor captures the shared genetic liability to suicidality, while residual components indicate genetic influences specific to each phenotype. Complicating the genetic architecture of suicidality phenotypes is their genetic correlations with psychiatric disorders. Thus, we extend these models to include 14 psychiatric disorders (bipolar disorder, schizophrenia, alcohol use disorder, cannabis use disorder, opioid use disorder, tobacco use disorder, attention-deficit/hyperactivity disorder (ADHD), autism spectrum disorder, Tourette’s syndrome, anorexia nervosa, obsessive compulsive disorder, anxiety, major depressive disorder, and PTSD), for which the shared genetic architecture and latent factor structure have been previously established in large-scale cross-disorder genomic SEM analyses^39^. This decomposes the genetic variance in suicidality into components shared with, and not fully accounted for by broader psychiatric liability. Such models can increase power for genetic discovery and provide a more nuanced interpretation of the genetic architecture underlying complex phenotypes^24,38–42^.

Here, we applied genomic SEM to GWAS of SI, SA, SD, and 14 psychiatric disorders to characterize their shared and distinct genetic architectures. We identified a suicidality common factor that captures shared genetic liability across SI, SA, and SD, alongside phenotype-specific genetic components, particularly for SD. We further demonstrated that both the suicidality common factor and individual suicidality phenotypes retain significant residual genetic variance after accounting for their shared genetics with psychiatric disorders. Together, these findings refine our understanding of the genetic architecture of suicidality phenotypes.

## RESULTS

### Suicidality common factor

The common factor model, which models the shared genetic covariance among suicidality phenotypes as a single latent factor, showed high loadings of SI, SA, and SD onto a latent suicidality common factor (Fig. 1). SA exhibited the highest loading, whereas SD showed the lowest loading and the largest residual variance. The three-indicator common factor model is just-identified (i.e., it has zero degrees of freedom), meaning it perfectly reproduces the observed genetic covariance matrix (𝑋^2^(0)=0, comparative fit index (CFI)=1.00, standardized root mean square residual (SRMR)=0); therefore, global fit indices for the suicidality common factor model do not provide meaningful information about model quality and were not used to assess model fit in this instance. We also tested an alternative model that included NSSI^16^ as a fourth indicator (Supplementary Fig. 1). Although this model fit the data well (𝑋^2^(2)=0.21, CFI=1.00, SRMR=0.016), our primary interest was the genetic architecture of suicidality specifically, rather than broader self-injurious behavior. Consistent with this distinction, NSSI showed a comparatively weaker loading (0.56) and larger residual variance, indicating that it shared less genetic covariance with the suicidality indicators. Thus, we retained the more parsimonious three-indicator suicidality common factor model for downstream analyses (Fig. 1).

**Figure 1.**
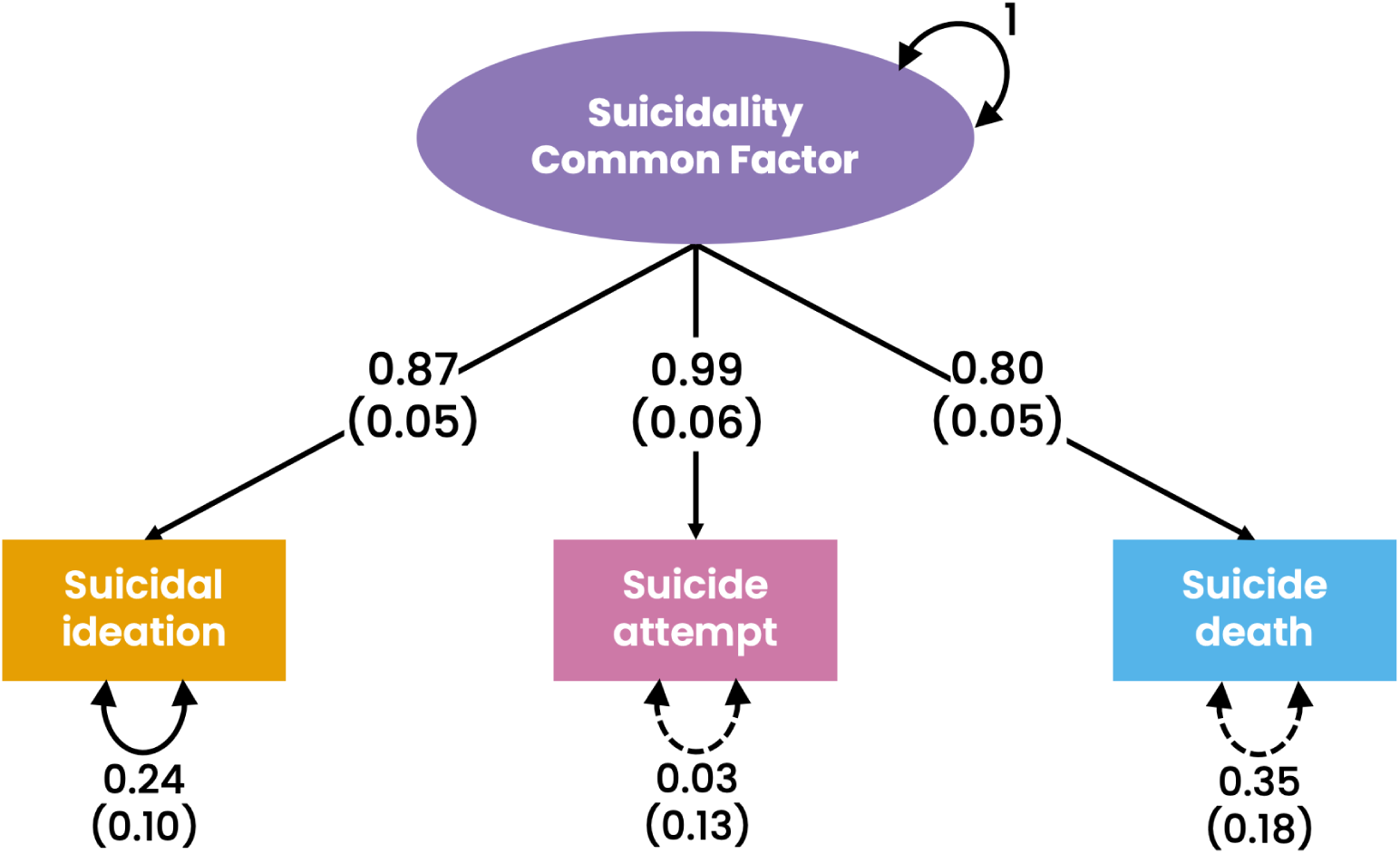
Suicidality common factor model. Path diagram for a model in which suicidal ideation, suicide attempt, and suicide death load on a latent suicidality common factor. Values on single-headed arrows represent standardized factor loadings and values on double-headed arrows represent residual variances. Standard errors are presented in parentheses. Solid lines represent significance at p<0.05 and dashed lines represent non-significance.

Next, we performed a common factor GWAS to estimate the effect of SNPs on the suicidality common factor. The GWAS identified 36 independent genome-wide significant (p< 5×10^-8^) loci (Fig. 2, Supplementary Table 1) and the mean 𝑋^2^ = 1.55 across variants used in LDSC. Seven out of these 36 loci were not previously reported as genome-wide significant in the contributing univariate GWAS (SI, SA, SD)^18^ or other previous suicidality GWAS^18,20,22,23,25–27,43,44^ (Fig. 2, Supplementary Table 1). To the best of our knowledge, those seven loci have also not been identified in previous psychiatric disorder GWAS (based on GWAS data deposited at GWAS catalog as of April 2026: https://www.ebi.ac.uk/gwas/home). Eight variants had significant heterogeneity (Q_SNP_) estimates, indicating effects not fully mediated by the common factor. These mapped to a chromosome 1 locus (lead SNP rs2274700) in the *CFH* gene and a chromosome 10 locus (lead SNP rs36212732) in the *ARMS2* gene (Fig. 2, Supplementary Table 2), the latter of which was also identified in the univariate SD GWAS^18^. Inspection of test statistics and p-values from the univariate GWAS suggested that both Q_SNP_ loci are likely driven by their effects specific to SD and not associated with SI or SA.

**Figure 2.**
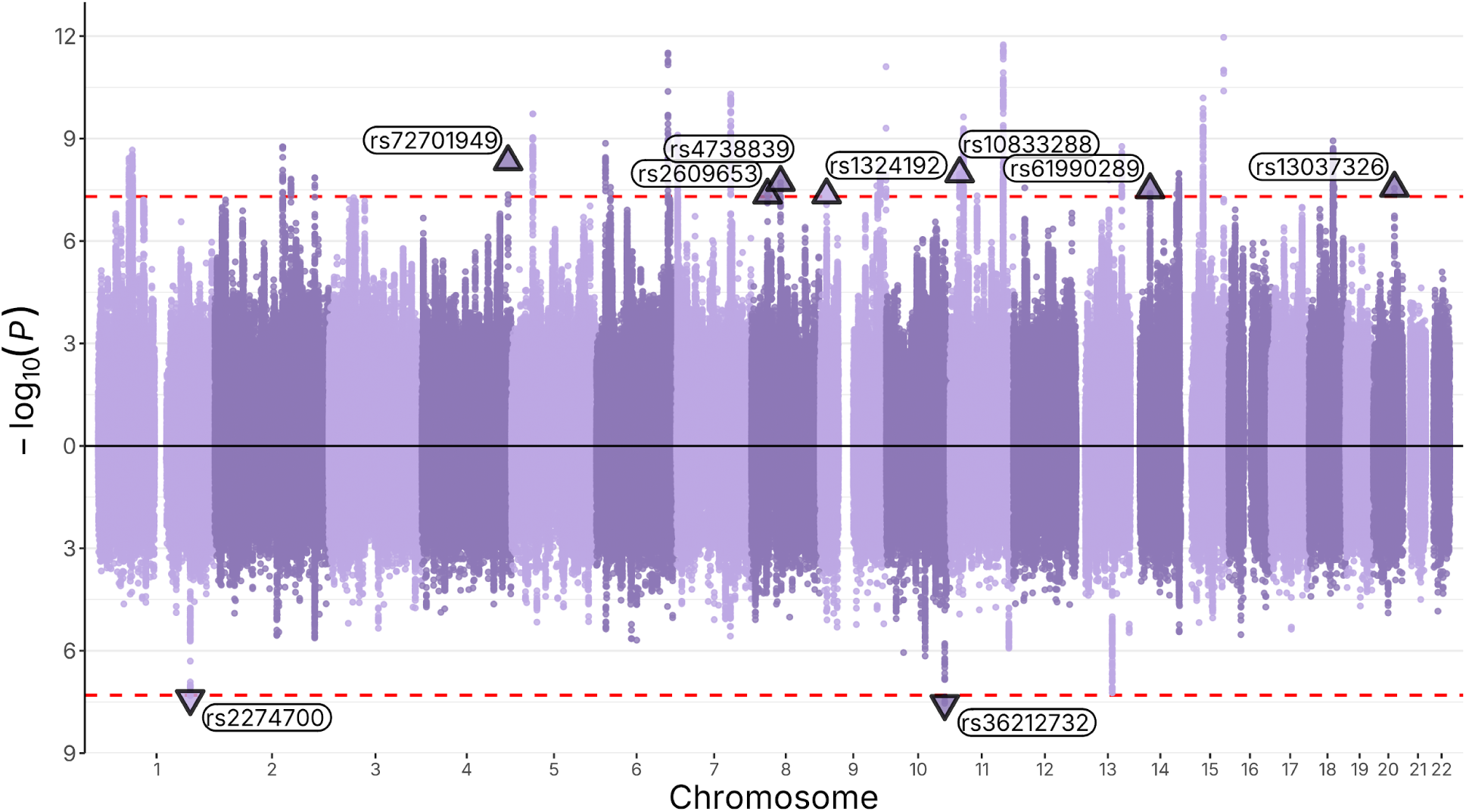
Miami plot of the suicidality common factor GWAS and Q_SNP_ estimates. The x-axis shows chromosomal position and the y-axis shows significance of the association as -log10(P). In the top half, purple points indicate associations with the suicidality common factor and purple triangles are lead SNPs from loci not previously reported as genome-wide significant in any of the univariate or previous suicidality GWAS. In the bottom half, purple points indicate significance of Q_SNP_ estimates and purple triangles are lead SNPs from the two significant Q_SNP_ loci. The red lines show the genome-wide significance threshold (p = 5×10^-8^).

### Phenotype-specific GWAS of SI, SA, and SD

We used the GWAS-by-subtraction method to perform SI-, SA-, and SD-specific GWAS to identify loci that contribute specifically to each phenotype (Supplementary Fig. 2). No loci reached genome-wide significance for the SI- or SA-specific GWAS (Supplementary Fig. 3). The SD-specific GWAS identified one genome-wide significant locus on chromosome 13 (lead SNP rs9318402; Supplementary Fig. 3C, Supplementary Table 3), which also had a near genome-wide significant Q_SNP_ estimate (p=5.92×10^-8^) in the common factor GWAS. Additionally, the two significant Q_SNP_ loci from the common factor GWAS were near genome-wide significant in the SD-specific GWAS (p<5×10^-7^).

### Local genetic correlations

Local genetic correlation analyses revealed ten, three, and one region(s) with significant positive genetic covariance between SI-SA, SI-SD, and SA-SD, respectively (Supplementary Table 4). One of these regions on chromosome 17 (hg19 coordinate: 65,016,683-66,315,468) showed significant positive genetic covariance between all three phenotype pairs, while all other regions were unique to either SI-SA or SI-SD. Of the 12 unique regions, six, including the chromosome 17 region, have not been previously reported as significantly (p<5×10^-8^) associated with suicidality phenotypes in the univariate^18^, common factor, or other previous suicidality GWAS^18,20,22,23,25–27,43,44^, but all were near genome-wide significant (all p<5.04×10^-6^) in the common factor GWAS.

### Conditioning suicidality phenotypes on psychiatric factors

Using four separate models, we regressed each of the original individual suicidality phenotypes (SI, SA, SD) and the suicidality common factor on five latent psychiatric factors constructed from GWAS of 14 psychiatric disorders (Table 1, Supplementary Figs. 4-7). The structure of the five latent psychiatric factors is based on the best-fitting model reported by Grotzinger et al. (2026)^39^. After conditioning on all five latent factors, final models were run in which only significant factors were retained (Fig. 3). For example, the compulsive factor was not significantly associated with SI, SA, SD or the suicidality common factor (Supplementary Figures 4-7) and therefore was removed. There was significant residual variance for SI, SA, SD and the suicidality common factor after conditioning on significant psychiatric factors, although it varied. SD showed the most residual variance (58%, Fig. 3C), SI the second most (23%, Fig. 3A), with the smallest residual variances (both 14%) observed for SA (Fig. 3B) and the suicidality common factor (Fig. 3D). The SCZ-BD factor (comprising schizophrenia and bipolar disorder) had a significant loading for all suicidality phenotypes (Fig. 3). For SI, the internalizing factor (INTERN; comprising anxiety, major depressive disorder, and PTSD) was the only other factor with a significant loading (Fig. 3A). For SD, the substance use disorder factor (SUD; comprising alcohol, cannabis, opioid, and tobacco use disorder and ADHD) was the only other factor with a significant loading (Fig. 3C). For SA and the suicidality common factor, SCZ-BD, INTERN, SUD and the neurodevelopmental factor (NEURO; comprising ADHD, autism spectrum disorder, and Tourette’s Syndrome) had significant loadings (Fig. 3B, D).

**Figure 3.**
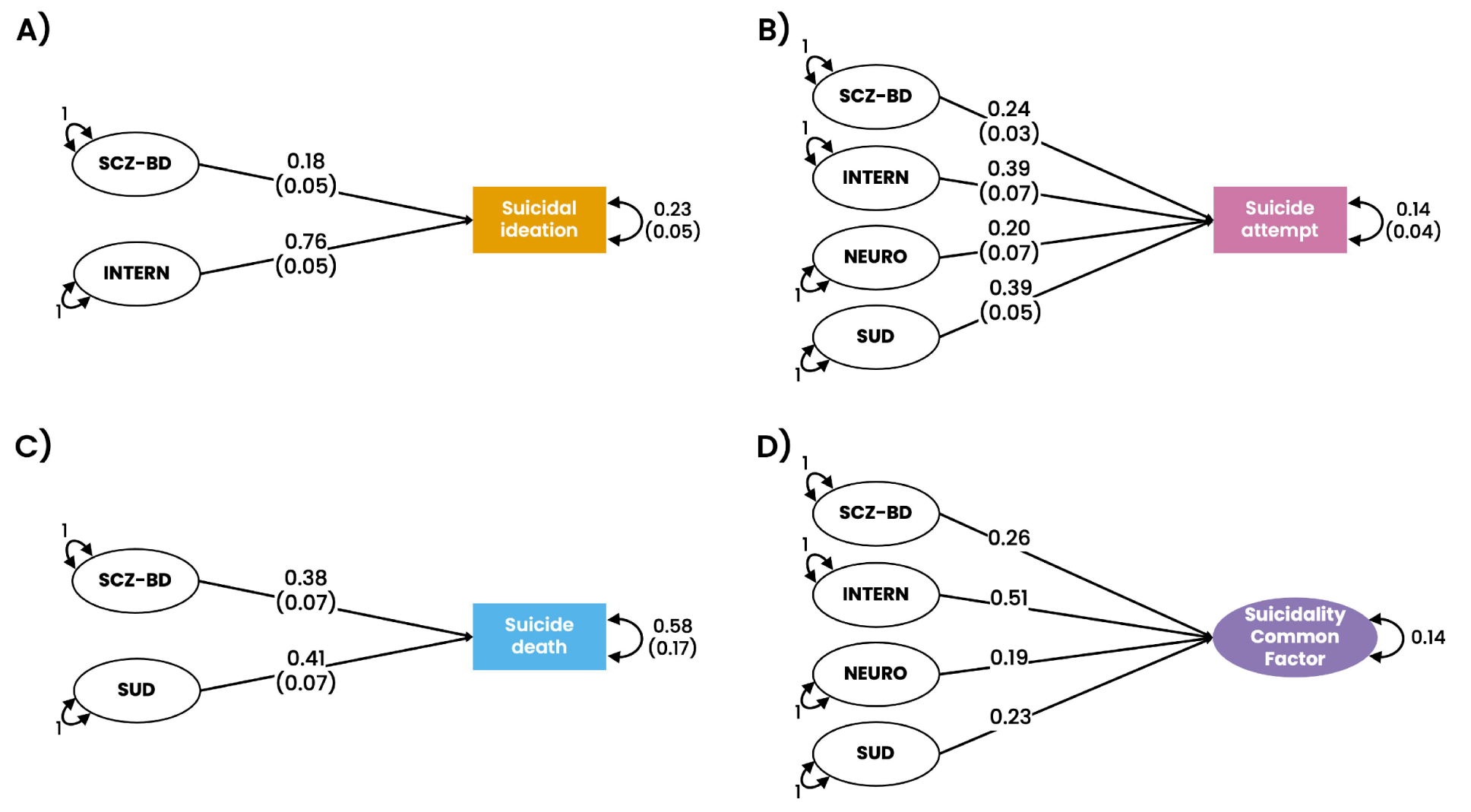
Path diagrams showing final models of suicidality phenotypes conditioned on significant psychiatric factors. A) Suicidal ideation conditioned on SCZ-BD (the schizophrenia-bipolar disorder factor) and INTERN (the internalizing factor). B) Suicide attempt conditioned on SCZ-BD, INTERN, NEURO (the neurodevelopmental factor), and SUD (the substance use disorder factor). C) Suicide death conditioned on SCZ-BD and SUD. D) The suicidality common factor conditioned on SCZ-BD, INTERN, NEURO, and SUD. For simplicity, indicators loading on to latent factors and the factor covariances are not shown, but provided in Supplementary Figures 4-7. Paths show loadings, residual variances, and their standard errors in parentheses. Standard errors are not shown for model D as the suicidality common factor is both a latent and endogenous variable in the model and therefore stable standard error estimates could not be obtained.

**Table 1:**
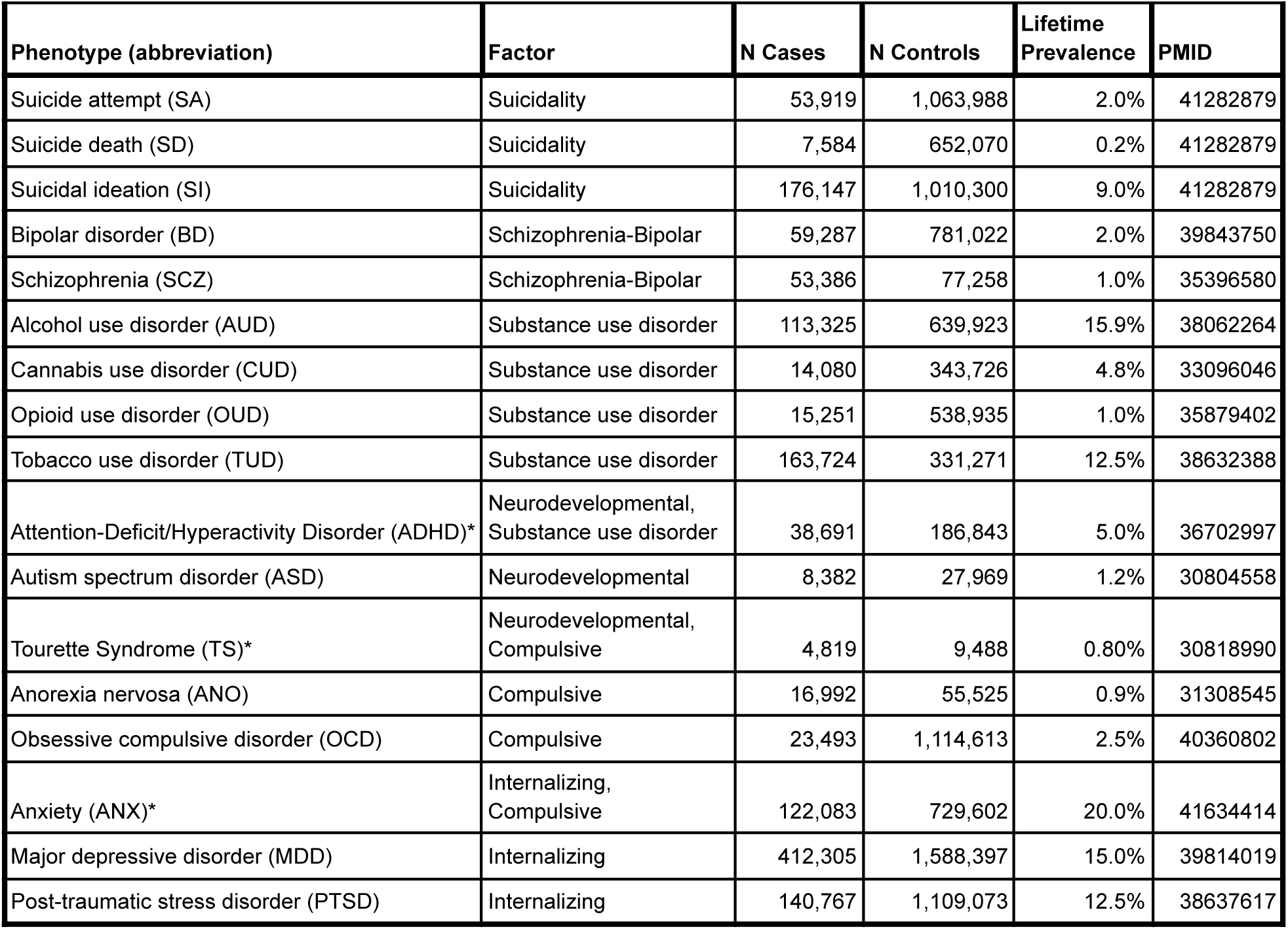
Descriptive statistics for the suicidality and psychiatric disorder univariate GWAS. The factor column lists the latent factor each individual phenotype loaded onto. *In some cases, individual phenotypes had cross-loadings on multiple latent psychiatric factors, following the best-fitting model structure from Grotzinger et al. (2026).

We next incorporated SNP effects into these models to perform GWAS of SI, SA, SD and the suicidality common factor conditional on significant psychiatric factors (Fig. 4, Supplementary Tables 5-8, Supplementary Figs. 8-11). No loci reached genome-wide significance in the conditional GWAS of SI, SA, or the suicidality common factor. One locus on chromosome 13 (lead SNP rs7335526) reached genome-wide significance for SD (Fig. 4C, Supplementary Table 7, Supplementary Fig. 10). We then evaluated whether genome-wide significant loci from the original GWAS were associated after conditioning on psychiatric disorders using a replication significance threshold (p = 0.05/number of genome-wide significant loci in the original GWAS). For SI, nine of 10 loci that were genome-wide significant in the univariate SI GWAS reached replication significance in the conditional SI GWAS (Fig. 4A, Supplementary Table 5, Supplementary Fig. 8); the remaining locus could not be tested because it was not present in the conditional GWAS (Supplementary Table 5). Of the 35 genome-wide significant loci identified in the univariate SA GWAS, 24 reached replication significance in the conditional GWAS (Fig. 4B, Supplementary Table 6, Supplementary Fig. 9). For SD, both previously identified genome-wide significant loci remained significant in the conditional SD GWAS (Fig. 4C, Supplementary Table 7, Supplementary Fig. 10). Of the 36 genome-wide significant loci from the suicidality common factor GWAS, 20 reached replication significance in the conditional GWAS (Fig. 4D, Supplementary Table 8, Supplementary Fig. 11).

**Figure 4.**
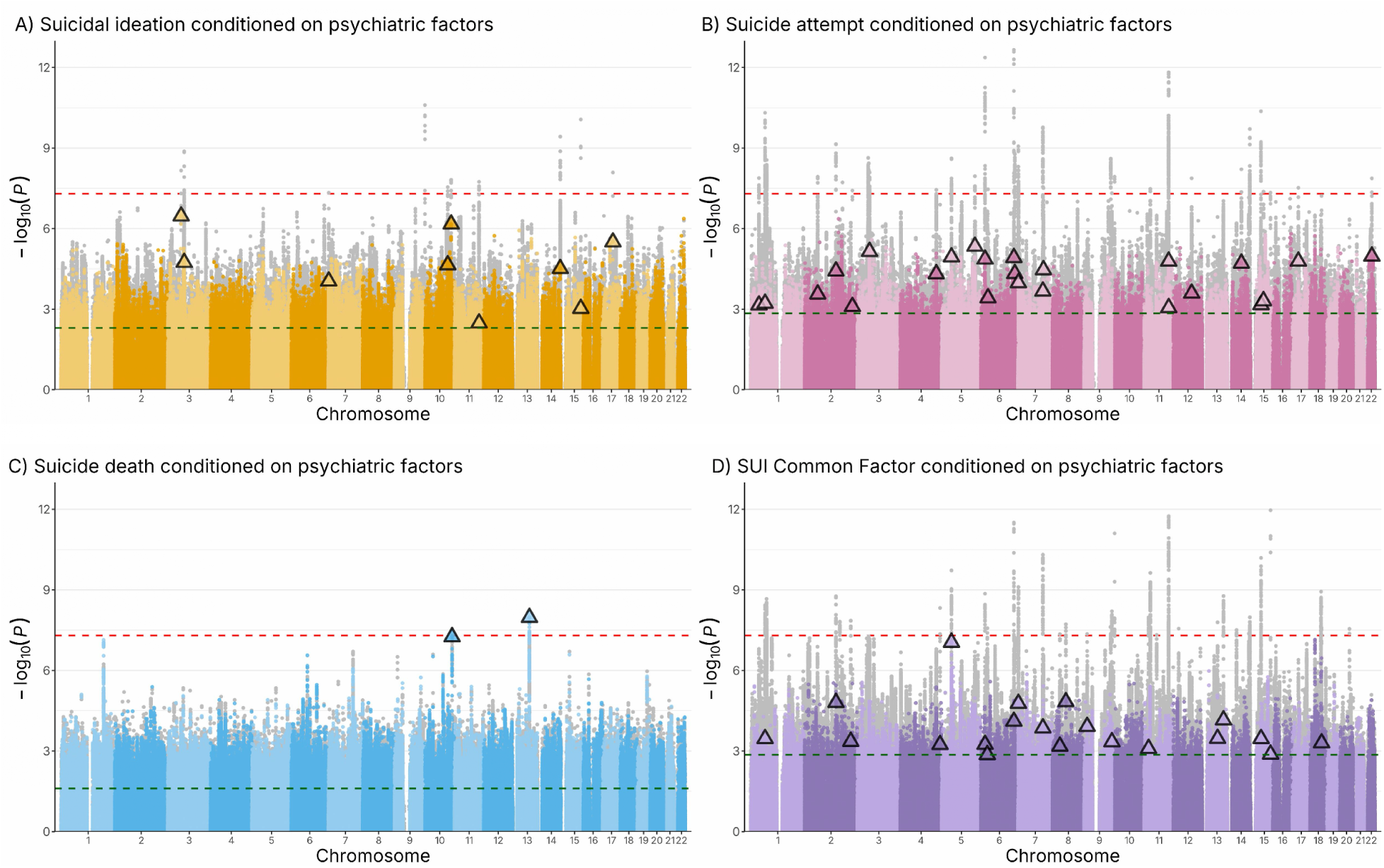
Manhattan plots of the GWAS of the suicidality phenotypes and suicidality common factor conditioned on latent psychiatric factors derived from GWAS of 14 psychiatric disorders. On each panel, the colored points show the GWAS after conditioning and the gray points show the original (i.e., univariate or common factor) GWAS. The x-axes show chromosomal position and the y-axes show significance of the association as -log10(P). The red lines show the genome-wide significance threshold (p = 5×10^-8^). The green lines show the replication significance threshold (p = 0.05/number of genome-wide significant loci in the univariate GWAS). Triangles indicate genome-wide significant loci from the original GWAS that reached replication significance after conditioning on the psychiatric factors. Forest plots showing the effect sizes and p-values before and after conditioning are in Supplementary Figures 8-11.

## DISCUSSION

We identified a latent suicidality common factor capturing substantial shared genetic variance across SI, SA, and SD, alongside phenotype-specific genetic components that differentiate individual phenotypes and were most prominent for SD. Our results support genetic contributions to suicidality that are independent of broader genetic liability to psychiatric disorders. Although this shared liability accounted for a meaningful proportion of variance across suicidality phenotypes, significant residual variance remained (12-58%), indicating that genetic liability to psychiatric disorders does not fully account for suicidality and supporting future examination of additional risk domains, such as physical health conditions^34,35,45^, impulsivity^46,47^, and loneliness^48,49^. Together, our findings provide evidence that phenotype-specific genetic variation distinguishes suicidality phenotypes beyond their shared liability.

The identification of the suicidality common factor supports the hypothesis that SI, SA, and SD share a core genetic liability, consistent with prior evidence of moderate-to-high genetic correlations across these phenotypes (0.70-0.88)^18,20^. Notably, SA showed the strongest loading on the suicidality common factor, consistent with the possibility that SA captures a broader constellation of features shared across suicidality phenotypes and may therefore more closely reflect their shared genetic liability. In this context, SA may represent an intermediate phenotype, with contributions from both internalizing and more behaviorally expressed (externalizing) factors, whereas SI and SD may each include greater phenotype-specific heterogeneity. For example, SI may index a relatively more internalizing form of psychopathology reflecting cognitive and affective processes (e.g., thoughts and rumination) rather than behavioral enactment^6–8,50^. SD showed a relatively weaker loading and the most residual variance, suggesting that while SD shares substantial genetic liability with other suicidality phenotypes, it may also be influenced by additional or distinct genetic or environmental factors^13,34,35,37,51,52^. This interpretation is further supported by our identification of three SD-specific loci according to Q_SNP_ and GWAS-by-subtraction results. These findings align with epidemiological and genetic studies suggesting that the progression from SI to SA to SD is not solely driven by increasing severity or an accumulation of environmental or polygenic risk for suicidality along a single continuum, but also involves partially distinct etiological pathways^37,53,54^. Furthermore, the modeling of a self-harm common factor showed that NSSI did not cluster with the suicidality phenotypes as well, in line with its weaker genetic correlations with SI, SA, and SD and the hypothesis that there may be a genetic component specific to suicidality and related to desire to die, rather than to only harm oneself^15–18^.

Our multivariate GWAS of the suicidality common factor identified 36 genome-wide significant loci, including seven not detected in prior univariate analyses, demonstrating increased power for locus discovery using this approach. At the same time, identification of phenotype-specific loci for SD highlights the importance of modeling heterogeneity across suicidality phenotypes. The chromosome 13 locus (lead SNPs rs9318402 and rs7335526), identified in the SD-specific GWAS and GWAS of SD conditioned on psychiatric factors, may be of particular interest as a potentially SD-specific signal. This locus maps to an intergenic region near LINC00561 and, to the best of our knowledge, has not been implicated in any prior GWAS (based on GWAS data deposited at GWAS catalog as of April 2026: https://www.ebi.ac.uk/gwas/home), highlighting it as a potential candidate for further investigation. The Q_SNP_ signals at the *CFH* and *ARMS2* loci, and their convergence with SD-specific GWAS results, provide evidence for genetic effects that are not shared with SI or SA. *CFH* encodes complement factor H, which protects host cells against the complement response, thereby regulating the immune system^55^. The function of *ARMS2* (Age-Related Maculopathy Susceptibility 2) is not firmly established, although it may be involved in modulating oxidative stress^56^. Local genetic correlation analyses further demonstrated that shared genetic influences across suicidality phenotypes are not uniformly distributed across the genome but instead cluster within specific loci. The greater number of loci showing overlap between SI and SA than SD is consistent with greater genetic similarity between SI and SA, while SD showed more pronounced phenotype-specific residual variance.

By incorporating psychiatric disorders into our models, we assessed the extent to which suicidality reflects shared versus independent genetic liability with broader psychopathology. Although all suicidality phenotypes showed significant genetic overlap with psychiatric factors, substantial residual genetic variance remained after conditioning, particularly for SD. This indicates that suicidality is not fully explained by liability to psychiatric disorders and supports the existence of independent genetic contributions^11,57,58^. Moreover, the pattern of associations with psychiatric factors differed across phenotypes, with internalizing liability more strongly related to SI, substance use disorder-related liability to SD, and broader cross-disorder liability contributing to SA and the suicidality common factor. The internalizing factor showed the largest loading on the suicidality common factor and SI, and was among the strongest contributors to SA, but did not significantly load on SD, suggesting that internalizing liability primarily contributes to general and earlier-stage suicidality risk rather than to SD. The association between SD and SUD-related liability is particularly notable and may partly reflect underlying behavioral dimensions such as impulsivity, which are only partially captured by the SUD factor (indexed by SUDs and ADHD), but hypothesized to contribute to the transition from ideation to action^6–9,59,60^. These differences underscore the heterogeneity of suicidality and suggest that distinct clusters of psychiatric risk may contribute to different stages or manifestations^29,61–64^. Consistent with this interpretation, conditional GWAS analyses demonstrated that a substantial proportion of genome-wide significant loci identified in the original GWAS remained associated at a replication significance threshold after conditioning on latent psychiatric factors. These findings indicate that a portion of the genetic signal underlying suicidality is not fully explained by genetic liability shared with psychiatric disorders^11,20,57,58^.

Several limitations should be considered. First, our analyses were restricted to GWAS in individuals of European ancestry, due to the lack of sufficiently powered or available GWAS of suicidality phenotypes in other genetic ancestries^18^, which limits our ability to determine if these findings are applicable to other populations. Second, while sufficiently powered for genetic correlation and genomic SEM analyses, the SD GWAS is not as large or well powered as the GWAS for other phenotypes and SD cases were primarily derived from a single cohort^25^. This restricts any follow-up of the SD-specific associations discovered here, until additional external cohorts are available to replicate discoveries. Finally, we limited our analyses to 14 psychiatric disorders known to cluster onto correlated latent factors based on previous work^39^; however, other conditions, such as personality, behavioral, and sleep disorders, may also be relevant to suicidality. As sufficient data become available to model these conditions as latent constructs, it may be possible to examine their relationships with suicidality in future work.

## CONCLUSIONS

Our results demonstrate that suicidality reflects both a shared genetic liability across phenotypes and phenotype-specific genetic influences, with SD showing greater phenotype-specific genetic variance. Multivariate genomic SEM approaches, including a common factor analysis and GWAS-by-subtraction, enhanced locus discovery and provided a more nuanced understanding of the genetic architecture underlying suicidality and psychiatric phenotypes. These findings inform models of suicide risk and future genetic studies of suicidality, which should account for both shared and phenotype-specific genetic contributions.

## METHODS

### SAMPLES

We used the largest summary statistics from GWAS of three suicidality phenotypes and 14 psychiatric disorders (Table 1). Summary statistics for the suicidality phenotypes were obtained from the PGC SUI group’s most recent GWAS meta-analyses of SI, SA, and SD^18^.These GWAS were confirmed to be sufficiently powered for genomic SEM analyses, as all phenotypes demonstrated *h^2^_SNP_*Z scores > 5, following recommendations from the genomic SEM developers (https://groups.google.com/g/genomic-sem-users/c/Wn1jptU2VcY/m/9CWxfQbvAAAJ). The LDSC mean 𝑋^2^ = 1.40 for SI, 1.43 for SA, and 1.07 for SD. The psychiatric disorders and their corresponding GWAS used were based on previously validated models of genetic liability to psychiatric disorders from the PGC Cross Disorder working group and publicly available GWAS results^39^. All input and resulting GWAS were based on genomic coordinates aligned to the human genome build hg19 (GRCh37). For all phenotypes, we used summary statistics from GWAS generated using European-ancestry samples. As these GWAS represent meta-analyses, the sum of effective sample size across contributing cohorts is necessary to accurately calculate liability scale heritability of the phenotypes. Accordingly, if the sum of effective sample sizes was not provided in the summary statistics file, it was calculated using cohort sample size information from the corresponding reference; if that information was not available, it was computed using the method described in Grotzinger et al. (2023)^65^.

### CONFIRMATORY FACTOR ANALYSIS

We applied genomic structural equation modelling (genomic SEM)^38^ to model the shared genetic architecture of suicidality phenotypes. First, we used linkage disequilibrium score regression (LDSC)^66,67^ within the genomic SEM package to estimate the genetic covariance matrix among the suicidality phenotypes (SI, SA, and SD). We then fit a common factor model (using the “*commonfactor*” function) in which all three indicators loaded onto a single latent suicidality factor (SUI common factor). Factor loadings were freely estimated, and the variance of the latent factor was fixed to 1.0 to achieve model identification. Because the three-indicator suicidality common factor model was just-identified (i.e., zero degrees of freedom so the model fit the observed data perfectly), global fit indices were not informative; however, the estimated factor loadings indicate substantial shared genetic variance across suicidality phenotypes captured by the common factor. We also tested an alternative model of a self-harm common factor model that additionally included NSSI as a fourth indicator. Fit statistics could not be directly compared between the two models because the three-indicator suicidality common factor model is just-identified and, by definition, fits the observed data perfectly. As a result, fit indices for the suicidality common factor model are not informative, and model evaluation instead focused on parameter estimates and theoretical plausibility. In the four-indicator self-harm common factor model, NSSI showed a substantially weaker loading and large significant residual variance, indicating poorer alignment with the latent construct. Thus, including the less-closely-related NSSI indicator reduced parsimony without improving measurement of the construct, and we retained the three-indicator suicidality common factor as the primary model.

Next, we extended this framework to evaluate the extent to which the shared genetic liability to suicidality was independent of broader genetic liability to psychiatric disorders. To do so, we specified a series of structural regression models in which suicidality phenotypes (SI, SA, SD) or the suicidality common factor were conditioned on 14 psychiatric disorders. In these models, the relationships between the psychiatric disorders and their loadings onto broader latent psychiatric factors were specified following the model described by Grotzinger et al. (2026)^39^, and each suicidality phenotype (or the latent suicidality common factor) was regressed on these psychiatric factors simultaneously. This approach allowed for the estimation of residual genetic variance in suicidality after accounting for shared genetic influences with psychiatric disorders, while also estimating the unique contribution of each psychiatric factor to suicidality. Psychiatric factors were allowed to covary with one another, and all parameters were estimated jointly within the genomic SEM framework. The final conditional models only included psychiatric domains that had significant associations with the specific suicidality phenotype of interest.

### MULTIVARIATE GWAS

Genomic SEM performs multivariate GWAS by incorporating SNP effects from each input GWAS into the genetic covariance matrix, then fitting a model in which SNP effects are included at the level of the latent factor(s). Multivariate GWAS in this study were conducted using two approaches, depending on the model structure. First, the GWAS of the suicidality factor was conducted using the “*commonfactorGWAS*” function in genomic SEM, in which the SNP effects from each input GWAS are incorporated into the genetic covariance matrix, and the effect of the SNP on the common factor is estimated. This approach requires that SNP effects are available across all input GWAS; therefore, analyses were restricted to variants present in every GWAS used in the specific model, with SNPs missing from any input GWAS excluded. This function also estimates a heterogeneity index (Q_SNP_), which can be used to identify variants that do not act entirely through the common factor, i.e., variants that show heterogeneity in their effects across suicidality phenotypes. Using a previously described formula^41^, we calculated the sample size for the suicidality common factor to be 547,614.

Second, we used the GWAS-by-subtraction method^42^ to characterize genetic influences unique to specific suicidality phenotypes using a series of conditional GWAS (Supplementary Figure 2). The GWAS-by-subtraction method applies a Cholesky decomposition to decompose the variance in the correlations amongst factors, resulting in a GWAS of the residual genetic variance in a phenotype after conditioning on other phenotypes. For example, the SD-specific GWAS captures genetic effects on SD that are independent of genetic influences shared with SI and SA. This framework was also used to obtain GWAS on the residual ‘non-psychiatric’ genetic variation in SI, SA, SD, and the suicidality common factor by conditioning them on latent psychiatric factors. For these GWAS, suicidality phenotypes were only conditioned on psychiatric factors which were significant in the conditional models before SNP effects were included.

When running the conditional models and GWAS, genomic SEM reported a warning that in some cases, the genetic covariance matrix was non-positive definite, and as a result, the software applied smoothing to obtain a positive covariance matrix to use in model estimation. Post-hoc analyses in which each indicator was removed from the model indicated that the warning arose due to collinearity between the cannabis and opioid use disorder (OUD) GWAS, as they are highly genetically correlated. Thus, we removed the OUD GWAS from the model and smoothing was no longer necessary. To assess whether the extent of smoothing was problematic, we compared test statistics for SNPs with excessive smoothing in the full GWAS to the test statistics from the GWAS with OUD removed. All test statistics (beta, standard errors, Z-scores, and p-values) were highly correlated (R^2^ > 0.99), indicating that the smoothing did not significantly bias the conditional GWAS results. Accordingly, in the main text we present results from the full model, including OUD, for which smoothing was applied.

### SUPERGNOVA

We used SUPERGNOVA^68^ to identify specific genomic regions with significant local genetic correlations between SI, SA, and SD. We used partition files provided with the SUPERGNOVA software (https://github.com/qlu-lab/SUPERGNOVA), based on the hg19 (GRCh37) genome build, which included a total of 2,353 approximately independent LD genomic regions, resulting in a total of 2,078 regions in which there were sufficient numbers of SNPs in the provided GWAS. To correct for the number of regions tested, we applied a Bonferroni-corrected significance threshold of p < 2.4 x 10^-5^ (0.05/2,078 regions). We also tested for significant local genetic correlations between SD and age-related macular degeneration in regions harboring the *CFH* and *ARMS2* loci identified by the Q_SNP_ analysis and used a Bonferroni-corrected significance threshold of p < 0.025 (0.05/2 regions).

## DATA AVAILABILITY

Summary statistics from GWAS conducted here excluding the Utah SD cohort will be made available by application through the Psychiatric Genomics Consortium data access portal on publication: (https://pgc.unc.edu/for-researchers/data-access-committee/data-access-portal/). Data from the Utah site will be available upon request from the administrative manager of the Utah Suicide Mortality Research Study (https://rge.utah.edu/usmrs.php) and will be contingent upon the permissions required by institutional regulations and state statutes.

## CODE AVAILABILITY

All software used is publicly available at the URLs or references cited.

## ACKNOWLEDGEMENTS

We thank the participants who donated their time, life experiences, and DNA to this research, and the clinical and scientific teams that worked with them.

This material is based upon work supported by the National Science Foundation Graduate Research Fellowship Program under Grant No. 1842169 (PI Colbert) and the National Institute of Mental Health R01 MH132733 (PI Mullins). The PGC is supported by grants to UCSD (R01MH124847), UNC (R01MH124871), MGH (R01MH124851), Mount Sinai School of Medicine (R01MH124839), Cardiff University (R01MH124873), Trinity College Dublin (R01MH124875) and Washington University St. Louis (R01DA054869). The content is solely the responsibility of the authors and does not necessarily represent the official views of the US National Institutes of Health.

We thank SURF (www.surf.nl) for the support in using the National Supercomputer Snellius. Statistical analyses were carried out on the NL Genetic Cluster Computer (http://www.geneticcluster.org) hosted by SURFsara and the Mount Sinai high performance computing cluster (http://hpc.mssm.edu), which is supported by the Office of Research Infrastructure of the National Institutes of Health (Grant Nos. <u>S10OD018522</u> and <u>S10OD026880</u>).

## REFERENCES

1. World Health Organization. Suicide worldwide in 2021: global health estimates. https://www.who.int/publications/i/item/9789240110069 (2025).

2. World Health Organization. Preventing suicide: A global imperative. https://www.who.int/publications/i/item/9789241564779 (2014).

3. GBD 2021 Suicide Collaborators. Global, regional, and national burden of suicide, 1990-2021: a systematic analysis for the Global Burden of Disease Study 2021. Lancet Public Health 10, e189–e202 (2025).

4. Substance Abuse and Mental Health Services Administration (SAMHSA), Center for Behavioral Health Statistics and Quality. 2023 National Survey on Drug Use and Health (NSDUH) Releases. https://www.samhsa.gov/data/data-we-collect/nsduh-national-survey-drug-use-and-health/national-releases/2023

5. Centers for Disease Control and Prevention. Suicide Data and Statistics. Suicide Prevention https://www.cdc.gov/suicide/facts/data.html (2025).

6. Klonsky, E. D., Saffer, B. Y. & Bryan, C. J. Ideation-to-action theories of suicide: a conceptual and empirical update. Curr. Opin. Psychol. 22, 38–43 (2018).

7. Klonsky, E. D. & May, A. M. The three-step theory (3ST): A new theory of suicide rooted in the ‘ideation-to-action’ framework. Int. J. Cogn. Ther. 8, 114–129 (2015).

8. Joiner, T. Why People Die by Suicide. (Harvard University Press, 2005).

9. Van Orden, K. A. et al. The interpersonal theory of suicide. Psychol. Rev. 117, 575–600 (2010).

10. Voracek, M. & Loibl, L. M. Genetics of suicide: a systematic review of twin studies. Wien. Klin. Wochenschr. 119, 463–475 (2007).

11. Fu, Q. et al. A twin study of genetic and environmental influences on suicidality in men. Psychol. Med. 32, 11–24 (2002).

12. Brent, D. A. & Mann, J. J. Family genetic studies, suicide, and suicidal behavior. Am. J. Med. Genet. C Semin. Med. Genet. 133C, 13–24 (2005).

13. Edwards, A. C. et al. On the genetic and environmental relationship between suicide attempt and death by suicide. Am. J. Psychiatry 178, 1060–1069 (2021).

14. Lim, K. X., Krebs, G., Rimfeld, K., Pingault, J.-B. & Rijsdijk, F. V. Investigating the genetic and environmental aetiologies of non-suicidal and suicidal self-harm: a twin study. Psychol. Med. 52, 1–11 (2021).

15. Maciejewski, D. F. et al. Overlapping genetic and environmental influences on nonsuicidal self-injury and suicidal ideation: different outcomes, same etiology?: Different outcomes, same etiology? JAMA Psychiatry 71, 699–705 (2014).

16. Colbert, S. M. C. et al. Polygenic contributions to suicidal thoughts and behaviors in a sample ascertained for alcohol use disorders. Complex Psychiatry 9, 11–23 (2023).

17. Edwards, A. C., Singh, M., Peterson, R. E., Webb, B. T. & Gentry, A. E. Associations between polygenic liability to psychopathology and non-suicidal versus suicidal self-injury. Am. J. Med. Genet. B Neuropsychiatr. Genet. 195, e32982 (2024).

18. Colbert, S. M. C., Psychiatric Genomics Consortium Suicide Working Group, Ruderfer, D., Docherty, A. R. & Mullins, N. Genome-wide association studies identify 77 loci for suicidality and provide novel biological insights. medRxiv 2025.10.22.25338076 (2025) doi:10.1101/2025.10.22.25338076.

19. Ruderfer, D. M. et al. Significant shared heritability underlies suicide attempt and clinically predicted probability of attempting suicide. Mol. Psychiatry 25, 2422–2430 (2020).

20. Mullins, N. et al. Dissecting the shared genetic architecture of suicide attempt, psychiatric disorders, and known risk factors. Biol. Psychiatry 91, 313–327 (2022).

21. Kim, M. J. et al. In-Depth characterization of the shared genetic architecture of suicide attempts with other major psychiatric disorders. Transl. Psychiatry 16, 130 (2026).

22. Strawbridge, R. J. et al. Identification of novel genome-wide associations for suicidality in UK Biobank, genetic correlation with psychiatric disorders and polygenic association with completed suicide. EBioMedicine 41, 517–525 (2019).

23. Ashley-Koch, A. E. et al. Genome-wide association study identifies four pan-ancestry loci for suicidal ideation in the Million Veteran Program. PLoS Genet. 19, e1010623 (2023).

24. Colbert, S. M. C. et al. Exploring the genetic overlap of suicide-related behaviors and substance use disorders. Am. J. Med. Genet. B Neuropsychiatr. Genet. 186, 445–455 (2021).

25. Docherty, A. R. et al. Genome-wide association study of suicide death and polygenic prediction of clinical antecedents. Am. J. Psychiatry 177, 917–927 (2020).

26. Docherty, A. R. et al. GWAS meta-analysis of suicide attempt: Identification of 12 genome-wide significant loci and implication of genetic risks for specific health factors. Am. J. Psychiatry 180, 723–738 (2023).

27. Kimbrel, N. A. et al. Identification of novel, replicable genetic risk loci for suicidal thoughts and behaviors among US military veterans. JAMA Psychiatry 80, 135–145 (2023).

28. Nock, M. K. et al. Cross-national prevalence and risk factors for suicidal ideation, plans and attempts. Br. J. Psychiatry 192, 98–105 (2008).

29. Colbert, S. M. C. et al. Distinguishing clinical and genetic risk factors for suicidal ideation and behavior in a diverse hospital population. Transl. Psychiatry 15, 63 (2025).

30. Kim, C. D. et al. Familial aggregation of suicidal behavior: a family study of male suicide completers from the general population. Am. J. Psychiatry 162, 1017–1019 (2005).

31. Klonsky, E. D. & May, A. M. Differentiating suicide attempters from suicide ideators: a critical frontier for suicidology research. Suicide Life Threat. Behav. 44, 1–5 (2014).

32. Brent, D. A., Bridge, J., Johnson, B. A. & Connolly, J. Suicidal behavior runs in families. A controlled family study of adolescent suicide victims. Arch. Gen. Psychiatry 53, 1145–1152 (1996).

33. Franklin, J. C. et al. Risk factors for suicidal thoughts and behaviors: A meta-analysis of 50 years of research. Psychol. Bull. 143, 187–232 (2017).

34. Coon, H. et al. Absence of nonfatal suicidal behavior preceding suicide death reveals differences in clinical risks. Psychiatry Res. 347, 116391 (2025).

35. Xiao, Y. et al. Decoding suicide decedent profiles and signs of suicidal intent using latent class analysis. JAMA Psychiatry 81, 595–605 (2024).

36. Sun, S. et al. Genetic architecture of suicidal ideation continuum: latent profile analysis of data using the million veteran program cohort. Mol. Psychiatry 31, 2264–2272 (2026).

37. Coon, H. et al. Genetic liabilities to neuropsychiatric conditions in suicide deaths with no prior suicidality. *JAMA Netw*. Open 8, e2538204 (2025).

38. Grotzinger, A. D. et al. Genomic structural equation modelling provides insights into the multivariate genetic architecture of complex traits. *Nat*. Hum. Behav. 3, 513–525 (2019).

39. Grotzinger, A. D. et al. Mapping the genetic landscape across 14 psychiatric disorders. Nature 649, 406–415 (2026).

40. Hatoum, A. S. et al. Multivariate genome-wide association meta-analysis of over 1 million subjects identifies loci underlying multiple substance use disorders. Nat. Ment. Health 1, 210–223 (2023).

41. Mallard, T. T. et al. Multivariate GWAS of psychiatric disorders and their cardinal symptoms reveal two dimensions of cross-cutting genetic liabilities. Cell Genom. 2, 100140 (2022).

42. Demange, P. A. et al. Investigating the genetic architecture of noncognitive skills using GWAS-by-subtraction. Nat. Genet. 53, 35–44 (2021).

43. Kimbrel, N. A. et al. A genome-wide association study of suicide attempts in the million veterans program identifies evidence of pan-ancestry and ancestry-specific risk loci. Mol. Psychiatry 27, 2264–2272 (2022).

44. Erlangsen, A. et al. Genetics of suicide attempts in individuals with and without mental disorders: a population-based genome-wide association study. Mol. Psychiatry 25, 2410–2421 (2020).

45. Fanelli, G. et al. Polygenic risk scores for neuropsychiatric, inflammatory, and cardio-metabolic traits highlight possible genetic overlap with suicide attempt and treatment-emergent suicidal ideation. Am. J. Med. Genet. B Neuropsychiatr. Genet. 189, 74–85 (2022).

46. Gvion, Y. & Apter, A. Aggression, impulsivity, and suicide behavior: a review of the literature. Arch. Suicide Res. 15, 93–112 (2011).

47. Oquendo, M. A. & Mann, J. J. The biology of impulsivity and suicidality. Psychiatr. Clin. North Am. 23, 11–25 (2000).

48. Stickley, A. & Koyanagi, A. Loneliness, common mental disorders and suicidal behavior: Findings from a general population survey. J. Affect. Disord. 197, 81–87 (2016).

49. McClelland, H., Evans, J. J., Nowland, R., Ferguson, E. & O’Connor, R. C. Loneliness as a predictor of suicidal ideation and behaviour: a systematic review and meta-analysis of prospective studies. J. Affect. Disord. 274, 880–896 (2020).

50. May, A. M. & Victor, S. E. From ideation to action: recent advances in understanding suicide capability. Curr. Opin. Psychol. 22, 1–6 (2018).

51. Kendler, K. S. et al. Genetic liability to suicide attempt, suicide death, and psychiatric and substance use disorders on the risk for suicide attempt and suicide death: a Swedish national study. Psychol. Med. 53, 1639–1648 (2023).

52. Kim, M. J., Galfalvy, H., Singh, T. & Mann, J. J. Polygenic risk, trait variables, and external stressors in fatal and nonfatal suicidal behavior. *JAMA Netw*. Open 9, e2554325 (2026).

53. Kendler, K. S., Ohlsson, H., Sundquist, J., Sundquist, K. & Edwards, A. C. The sources of parent-child transmission of risk for suicide attempt and deaths by suicide in Swedish national samples. Am. J. Psychiatry 177, 928–935 (2020).

54. Sveticic, J. & De Leo, D. The hypothesis of a continuum in suicidality: a discussion on its validity and practical implications. Ment. Illn. 4, e15 (2012).

55. Parente, R., Clark, S. J., Inforzato, A. & Day, A. J. Complement factor H in host defense and immune evasion. Cell. Mol. Life Sci. 74, 1605–1624 (2017).

56. Pan, Y. et al. Exploring the contribution of ARMS2 and HTRA1 genetic risk factors in age-related macular degeneration. Prog. Retin. Eye Res. 97, 101159 (2023).

57. Oquendo, M. A., Wall, M., Wang, S., Olfson, M. & Blanco, C. Lifetime suicide attempts in otherwise psychiatrically healthy individuals. JAMA Psychiatry 81, 572–578 (2024).

58. Stone, D. M. et al. Vital signs: Trends in state suicide rates - United States, 1999-2016 and circumstances contributing to suicide - 27 states, 2015. MMWR Morb. Mortal. Wkly. Rep. 67, 617–624 (2018).

59. O’connor, R. C. Towards an Integrated Motivational–Volitional Model of Suicidal Behaviour. in International Handbook of Suicide Prevention 181–198 (John Wiley & Sons, Ltd, 2011).

60. O’Connor, R. C. & Kirtley, O. J. The integrated motivational-volitional model of suicidal behaviour. Philos. Trans. R. Soc. Lond. B Biol. Sci. 373, 20170268 (2018).

61. Park, S. et al. Association between level of suicide risk, characteristics of suicide attempts, and mental disorders among suicide attempters. BMC Public Health 18, 477 (2018).

62. Sampasa-Kanyinga, H. et al. Nonmedical use of prescription opioids, psychological distress, and suicidality among adolescents. Soc. Psychiatry Psychiatr. Epidemiol. 56, 783–791 (2021).

63. Poorolajal, J., Haghtalab, T., Farhadi, M. & Darvishi, N. Substance use disorder and risk of suicidal ideation, suicide attempt and suicide death: a meta-analysis. J. Public Health (Oxf.) 38, e282–e291 (2016).

64. DeVylder, J. E., Lukens, E. P., Link, B. G. & Lieberman, J. A. Suicidal ideation and suicide attempts among adults with psychotic experiences: data from the Collaborative Psychiatric Epidemiology Surveys: Data from the collaborative psychiatric epidemiology surveys. JAMA Psychiatry 72, 219–225 (2015).

65. Grotzinger, A. D., Fuente, J. de la, Privé, F., Nivard, M. G. & Tucker-Drob, E. M. Pervasive downward bias in estimates of liability-scale heritability in genome-wide association study meta-analysis: A simple solution. Biol. Psychiatry 93, 29–36 (2023).

66. Bulik-Sullivan, B. K. et al. LD Score regression distinguishes confounding from polygenicity in genome-wide association studies. Nat. Genet. 47, 291–295 (2015).

67. Bulik-Sullivan, B. et al. An atlas of genetic correlations across human diseases and traits. Nat. Genet. 47, 1236–1241 (2015).

68. Zhang, Y. et al. SUPERGNOVA: local genetic correlation analysis reveals heterogeneous etiologic sharing of complex traits. Genome Biol. 22, 262 (2021).

