## Supplementary Information for "Suicidality phenotypes reflect both shared and distinct genetic factors"

### SUPPLEMENTARY FIGURES

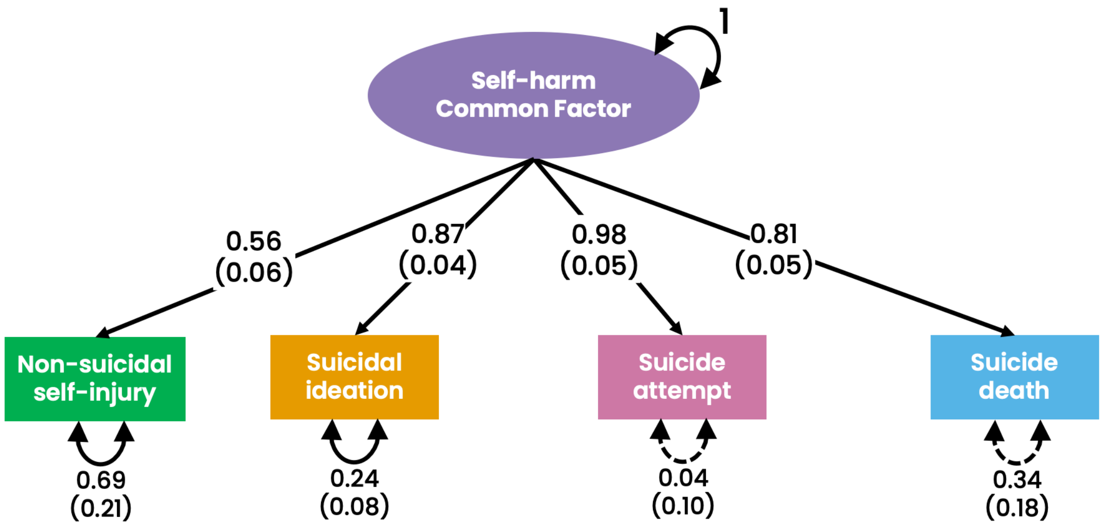

**Supplementary Figure 1. Alternative self-harm common factor model.** The path diagram for a model that allowed non-suicidal self-injury, suicidal ideation, suicide attempt, and suicide death to load on a latent self-harm common factor. Solid lines represent significance at p<0.05 and dashed lines represent non-significance. Standard errors are presented in parentheses. Model fit statistics: $X^2(2) = 0.21$, Akaike information criterion (AIC) = 16.21, comparative fit index (CFI) = 1.00, standardized root mean square residual (SRMR) = 0.016.

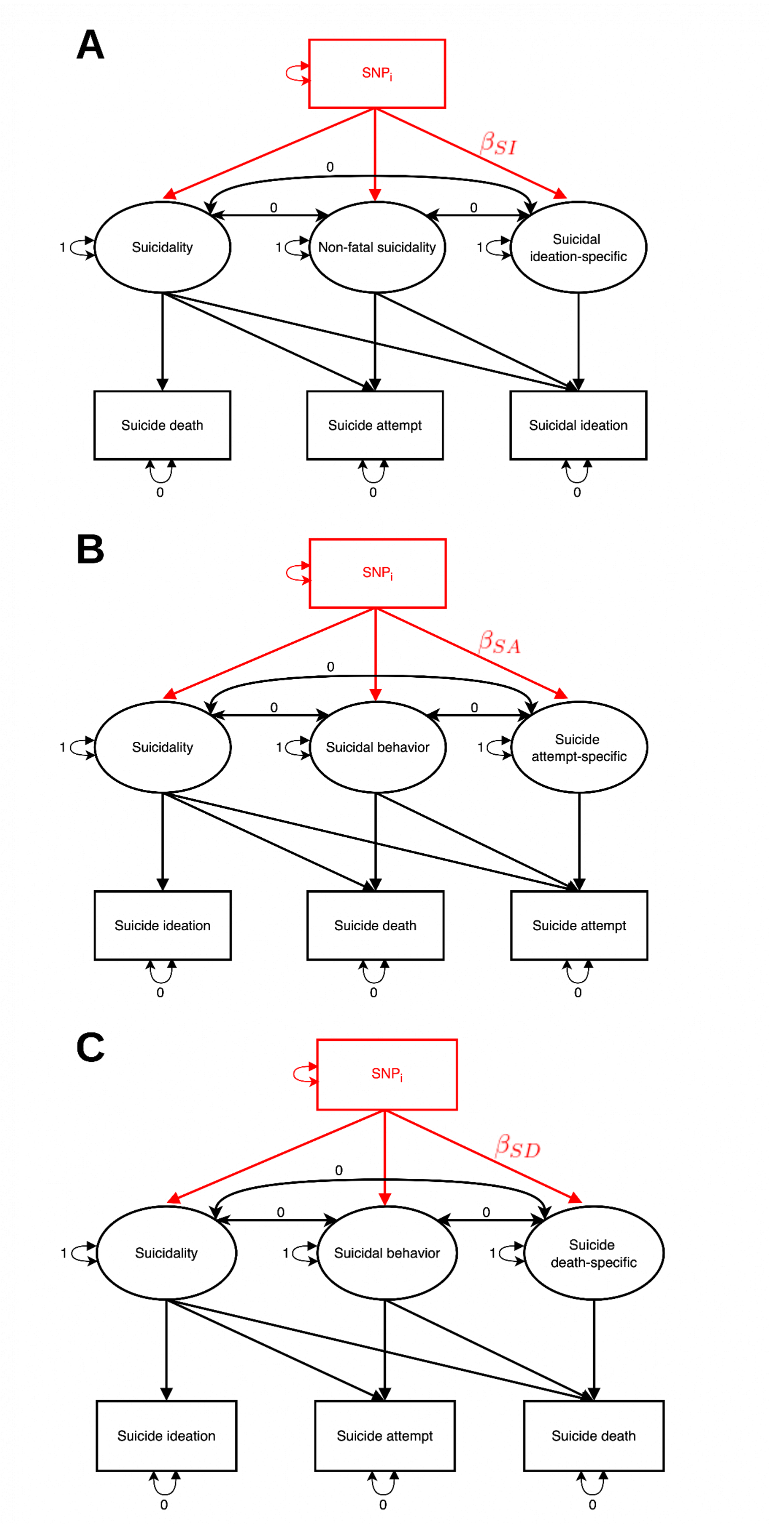

**Supplementary Figure 2. Cholesky decomposition and phenotype-specific GWAS models.** Black indicates the cholesky decomposition models with no SNP effects included and red indicates the models used for the phenotype-specific GWAS in which SNP effects on each cholesky factor are included. A) Cholesky decomposition of suicidal ideation-specific variance after accounting for covariance with suicide attempt and suicide death. B) Cholesky decomposition of suicide attempt-specific variance after accounting for covariance with suicidal ideation and suicide death. C) Cholesky decomposition of suicide death-specific variance after accounting for covariance with suicidal ideation and suicide attempt.

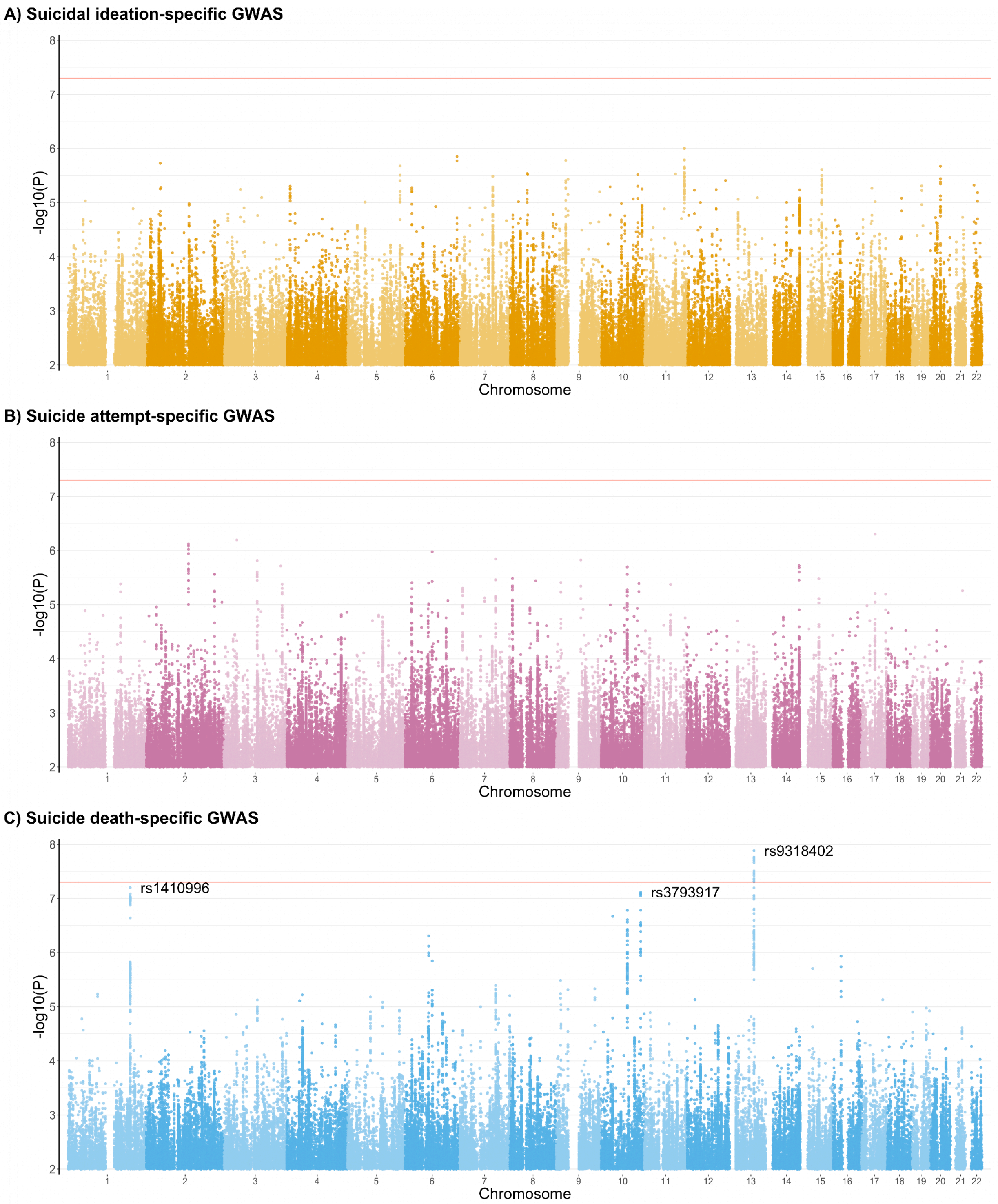

**Supplementary Figure 3. Manhattan plot of the suicidality phenotype-specific GWAS.** The x-axis shows chromosomal position and the y-axis shows significance of the association as -log10(P). The red line shows the genome-wide significance threshold ($p = 5\times10^{-8}$). A) Suicidal ideation-specific GWAS. B) Suicide attempt-specific GWAS. C) Suicide death-specific GWAS. Genome-wide significant and near-genome-wide significant loci are labelled with the lead SNP.

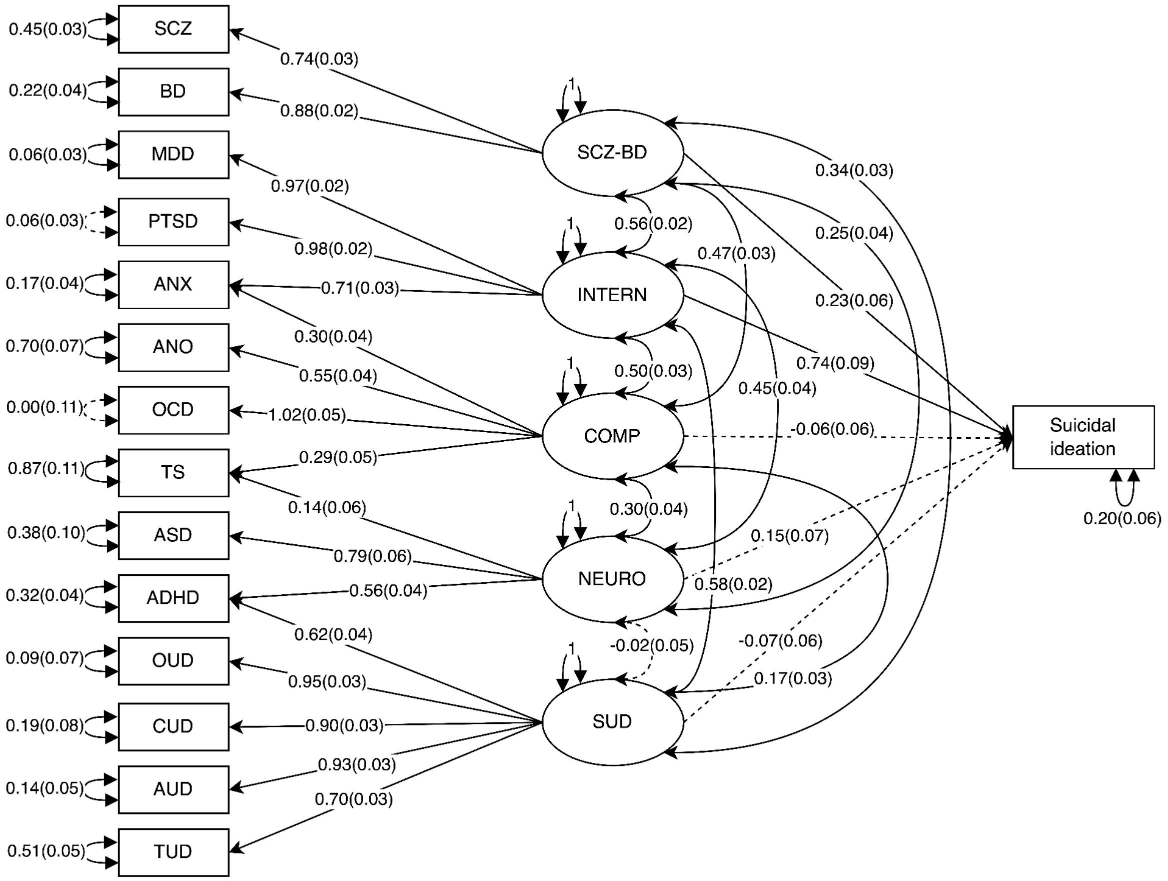

**Supplementary Figure 4. Full path diagram of suicidal ideation conditioned on all five latent psychiatric factors.** Paths show loadings, residual variances, covariances, and their standard errors in parentheses. Solid lines indicate significance ($p < 0.05$) while dashed indicates non-significance. The final model constructed based on these results is shown in the main text Fig. 3A. The 14 psychiatric disorders indicators are: SCZ= schizophrenia, BD = bipolar disorder, MDD = major depressive disorder, PTSD = post-traumatic stress disorder, ANX = anxiety, ANO = anorexia, OCD = obsessive compulsive disorder, TS = Tourette's syndrome, ASD = autism spectrum disorder, ADHD = Attention-Deficit/Hyperactivity Disorder, OUD = opioid use disorder, CUD = cannabis use disorder, AUD = alcohol use disorder, TUD = tobacco use disorder. The five latent psychiatric factors are: SCZ-BD = schizophrenia-bipolar disorder factor, INTERN = internalizing factor, COMP = compulsive factor, NEURO = neurodevelopmental factor, SUD = substance use disorder factor.

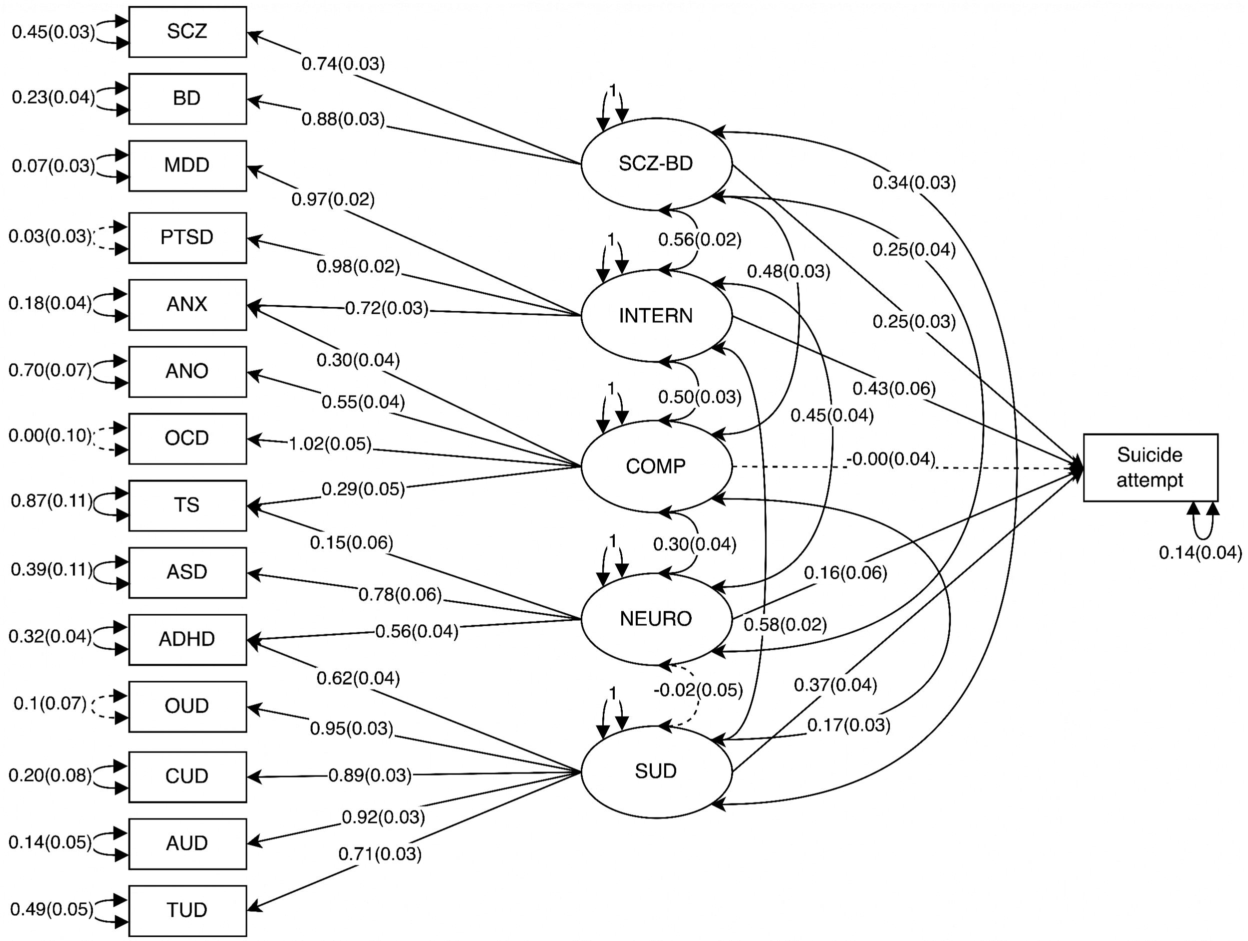

**Supplementary Figure 5. Full path diagram of suicide attempt conditioned on all five latent psychiatric factors.** Paths show loadings, residual variances, covariances, and their standard errors in parentheses. Solid lines indicate significance ($p < 0.05$) while dashed indicates non-significance. The final model constructed based on these results is shown in the main text Fig. 3B. The 14 psychiatric disorders indicators and five latent psychiatric factors are labelled the same as in Supplementary Figure 4.

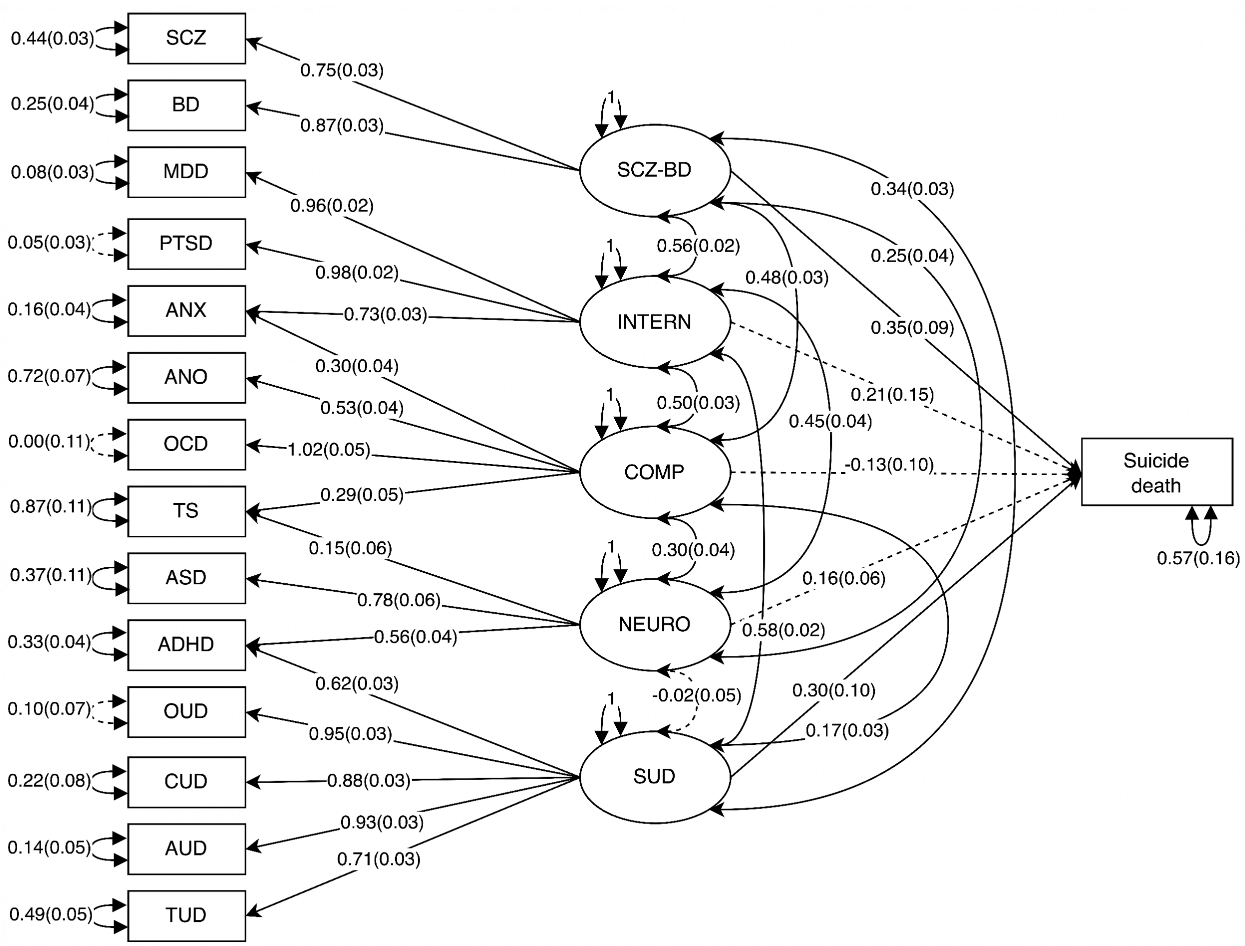

**Supplementary Figure 6. Full path diagram of suicide death conditioned on all five latent psychiatric factors.** Paths show loadings, residual variances, covariances, and their standard errors in parentheses. Solid lines indicate significance ($p < 0.05$) while dashed indicates non-significance. The final model constructed based on these results is shown in the main text Fig. 3C. The 14 psychiatric disorders indicators and five latent psychiatric factors are labelled the same as in Supplementary Figure 4.

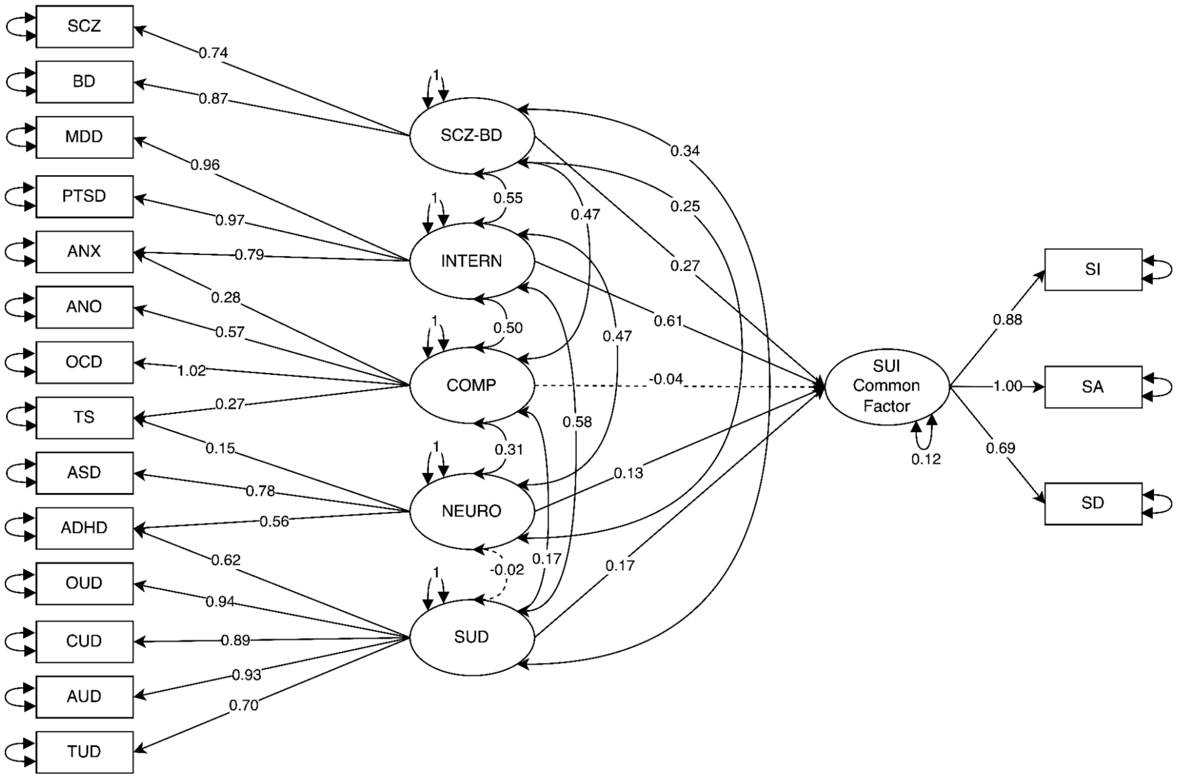

**Supplementary Figure 7. Full path diagram of the suicidality common factor conditioned on all five latent psychiatric factors.** Paths show loadings, residual variances, and covariances. Standard errors are not shown as the suicidality common factor is both a latent and endogenous variable in the model and therefore stable standard error estimates could not be obtained. Solid lines indicate significance ($p < 0.05$) while dashed indicates non-significance. The final model constructed based on these results is shown in the main text Fig. 3D. The 14 psychiatric disorders indicators and five latent psychiatric factors are labelled the same as in Supplementary Figure 4. SUI = suicidality, SI = suicidal ideation, SA + suicide attempt, SD = suicide death.

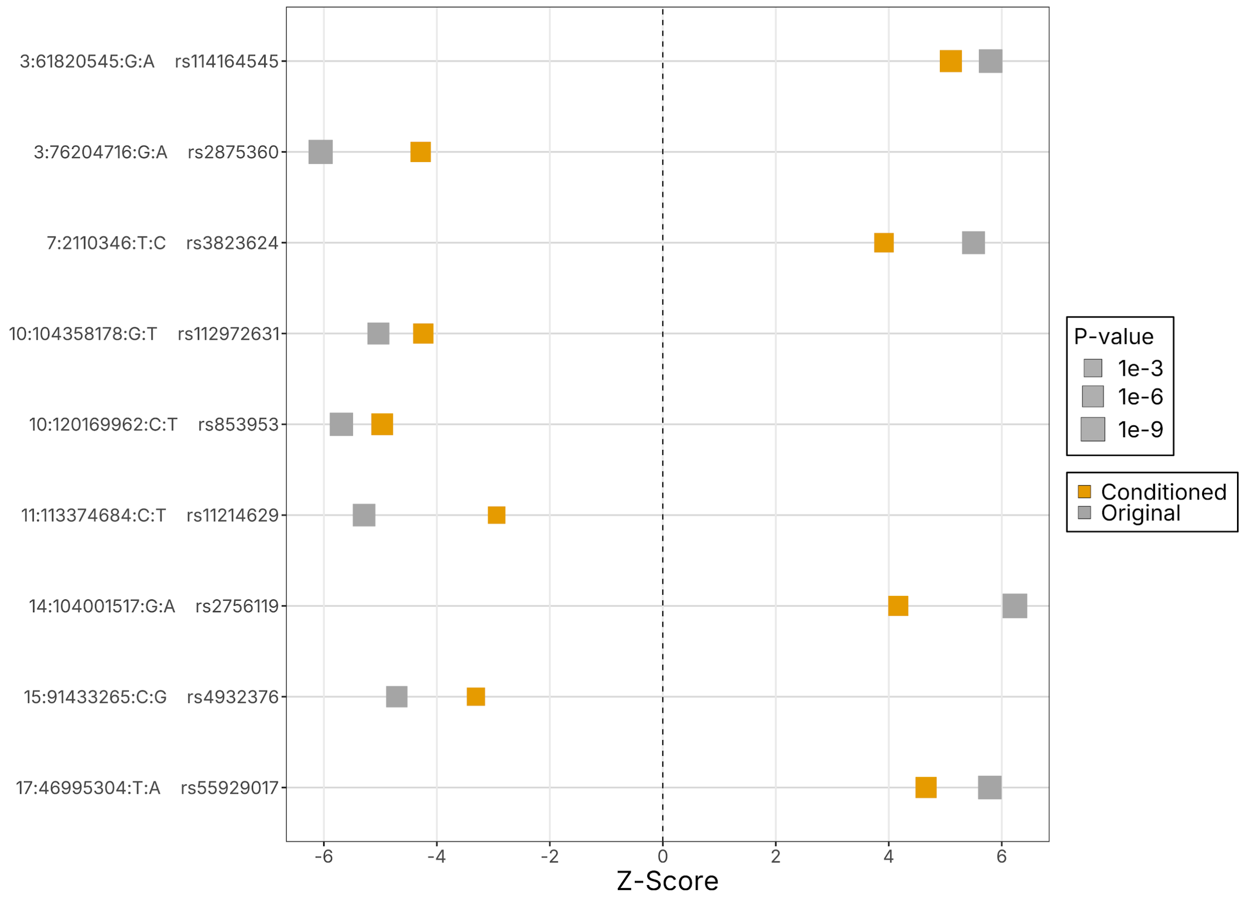

**Supplementary Figure 8. Forest plot of lead SNPs from loci significantly associated with suicidal ideation in the univariate GWAS.** Each box represents the Z-scores from either the GWAS of suicidal ideation conditioned on psychiatric factors (yellow) or the univariate suicidal ideation GWAS (gray). Box size corresponds to p-values, with size increasing as p-values become more significant.

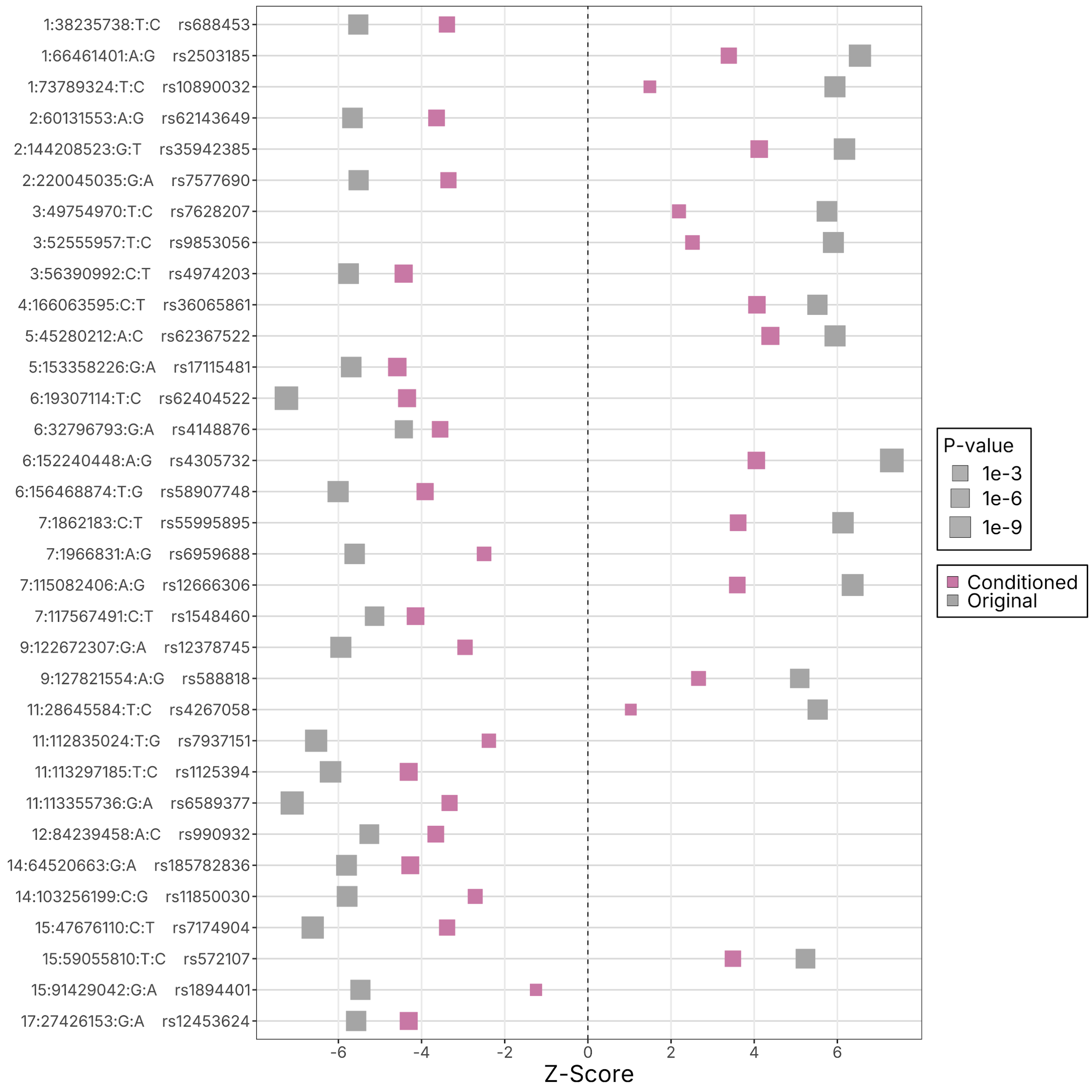

**Supplementary Figure 9. Forest plot of lead SNPs from loci significantly associated with suicide attempt in the univariate GWAS.** Each box represents the Z-scores from either the GWAS of suicide attempt conditioned on psychiatric factors (pink) or the univariate suicide attempt GWAS (gray). Box size corresponds to p-values, with size increasing as p-values become more significant.

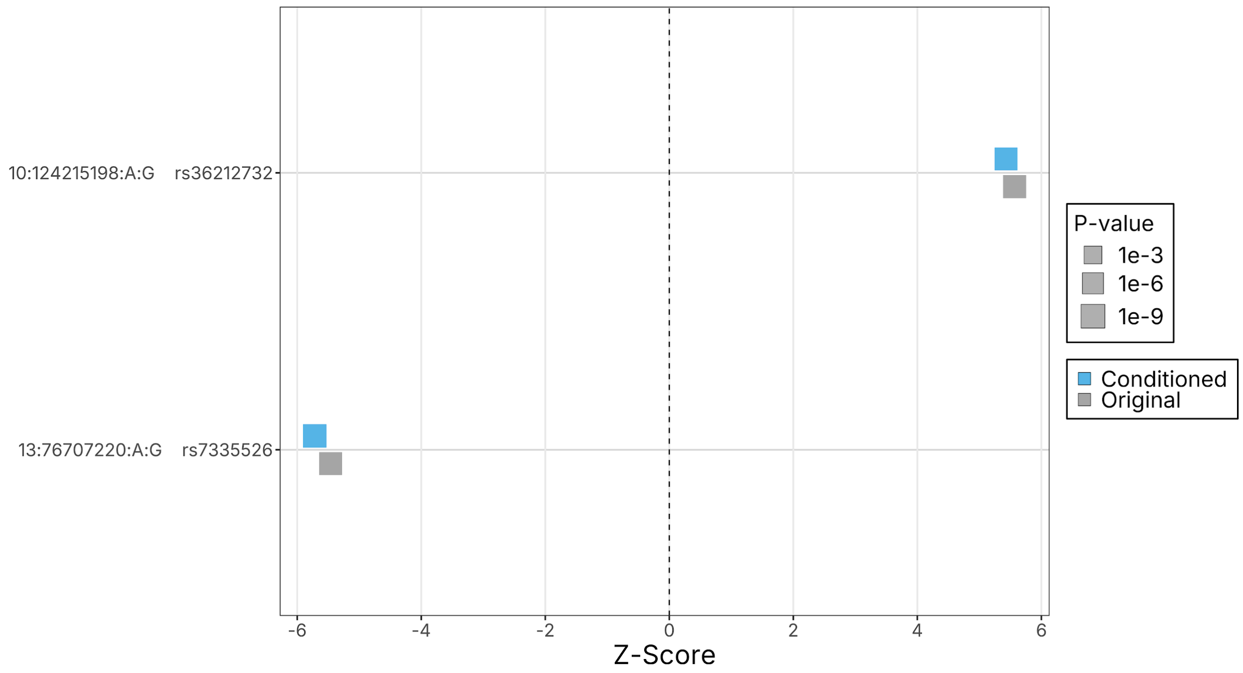

**Supplementary Figure 10. Forest plot of lead SNPs from loci significantly associated with suicide death in the univariate GWAS.** Each box represents the Z score from either the GWAS of suicide death conditioned on psychiatric factors (blue) or the univariate suicide death GWAS (gray). Box size corresponds to p-values, with size increasing as p-values become more significant.

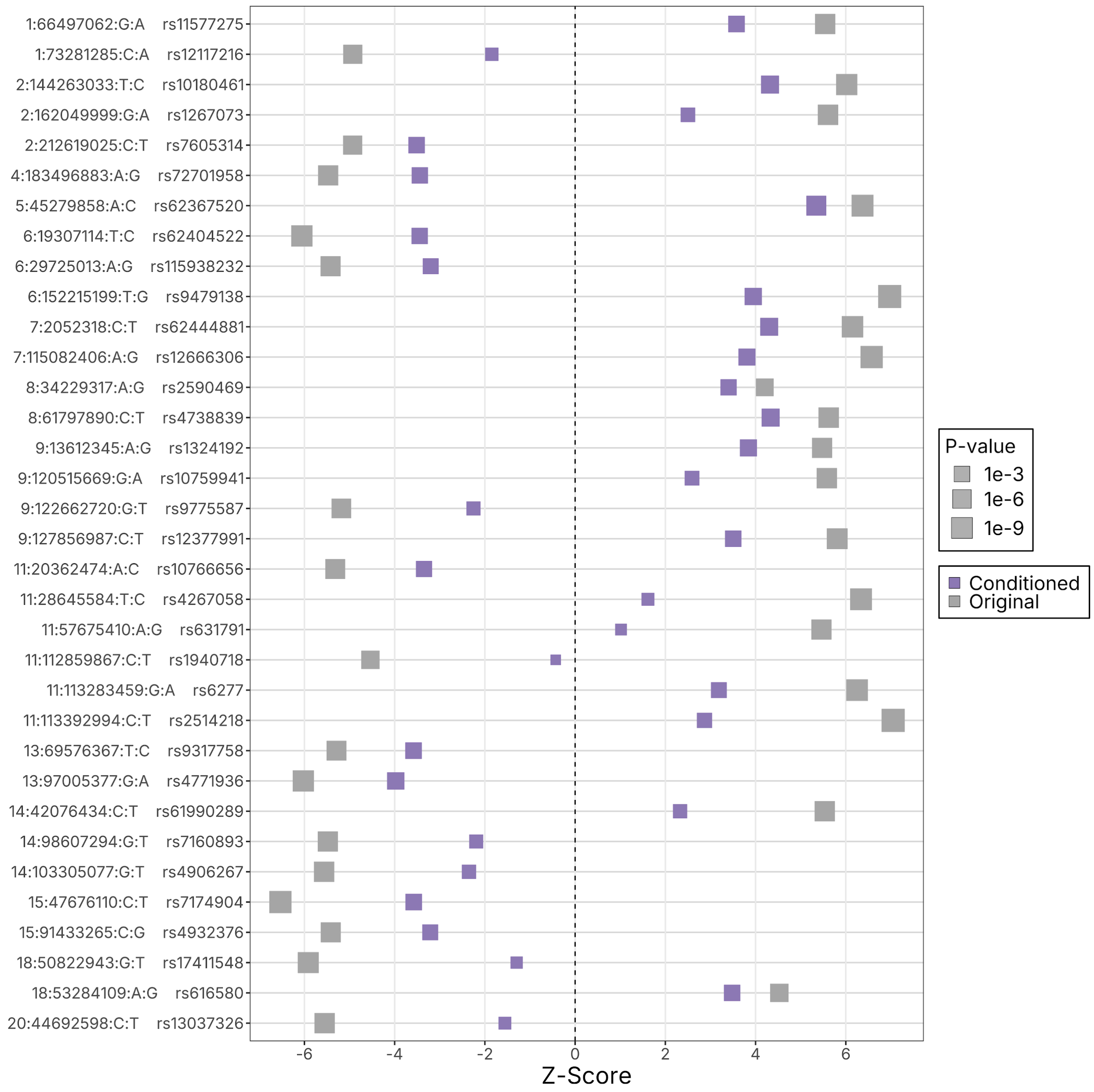

**Supplementary Figure 11. Forest plot of lead SNPs from loci significantly associated with the suicidality common factor.** Each box represents the Z-score from either the GWAS of the suicidality common factor conditioned on psychiatric factors (purple) or the primary suicidality common factor GWAS (gray). Box size corresponds to p-values, with size increasing as p-values become more significant.

### SUPPLEMENTARY NOTE

#### LIST OF CONSORTIUM MEMBERS FOR BYLINE CONSORTIUM AUTHORSHIP

**Members of the Suicide Working Group of the Psychiatric Genomics Consortium**

Sarah MC Colbert[1,2,3,4], Maria Koromina[2,3,1,4], Alexander S Hatoum[5,6], Mallory Stephenson[7,8], Alexis C Edwards[9,8], Emma C Johnson[5], Xuejun Qin[10,11], Andrey A Shabalin[12], Lucas T Ito[13,14,2,1,3], Kevin S O'Connell[15,16], Arvid Harder[17,18], Jens Hjerling-Leffler[17], Min Ji Kim[19], Ikuo Otsuka[20,21], Laura Vilar-Ribó[22], Arpana Agrawal[5], Martin Alda[23,24], Lars Alfredsson[25], Fazil Aliev[26,27], Till FM Andlauer[28], Celso Arango[29], Arnoud Arntz[30,31], Swapnil Awasthi[32,33], Olatunde O Ayinde[34], Silviu-Alin Bacanu[9], Peter B Barr[35,36,37], Claiton HD Bau[38,39], Bernhard T Baune[40,41], Jean C Beckham[42,43,44], Cosmin A Bejan[45], Sintia Belangero[46,14], Klaus Berger[47], Joanna M Biernacka[48,49], Ryan Bogdan[50], Dorret I Boomsma[51,52], Anders D Børglum[53,54,55], Kyle J Bourassa[56,57], David L Braff[22], Alice Braun[58,33], Rodrigo A Bressan[59,14,60], Tanja M Brueckl[61], Monika Budde[62], Brenda Cabrera-Mendoza[63,64], Bernardo Carpiniello[65], Zuriel Ceja[66,67], Jorge A Cervilla[68,69,70,71], Chiao-Erh Chang[72], Boris Chaumette[73,74,75], Myeong Jae Cheon[76], Sven Cichon[77,78,79], Jonathan RI Coleman[80,81,82], Hilary Coon[12,83,84,85], William E Copeland[86], Darina Czamara[61], Nina Dalkner[87], Udo Dannlowski[88,89], Friederike S David[90,91], Ditte Demontis[92,93], Arianna Di Florio[94], Carmen C Diaconu[95], Danielle M Dick[26,27], Dimitris Dikeos[96], Srdjan Djurovic[97,98], Howard J Edenberg[99,100], Annette Erlangsen[101,102,103,104], Sebastian Euler[105], Peter Falkai[106,107,108], Giuseppe Fanelli[109,110], Frederike T Fellendorf[87], Panagiotis Ferentinos[111], Fernando Fernandez-Aranda[112,113,114], Andreas J Forstner[90,115], Oleksandr Frei[116,117,118], Gabriel R Fries[119], Janice M Fullerton[120,121], Marie E Gaine[122,123,124], Hanga C Galfalvy[19,125], Marco Galimberti[63,126], Judith Garcia-Aymerich[127,128,129], Melanie E Garrett[130,11], Micha Gawlik[131,132], Joel Gelernter[133,134,135,136,137], Katherine Gordon-Smith[138], Aaron J Gorelik[50], Philip Gorwood[139,140], Hans J Grabe[141], Melissa J Green[142], Maria Grigoroiu-Serbanescu[143], Priya Gupta[144,145], Blanca Gutiérrez[68,69,146], Jose Guzman-Parra[147], Seonggyun Han[12], Marit Haram[117,148], Elizabeth R Hauser[10,11], Urs Heilbronner[62], Sabine C Herpertz[149], Jesús Herrera-Imbroda[147], Victor M Hesselbrock[150], Akitoyo Hishimoto[20], Bharath Holla[151], Anastasia Izotova[152,153,154], Yoonjeong Jang[155,156], Susana Jimenez-Murcia[112,114,113], Lisa A Jones[157], Lina Jonsson[158], JooEun Kang[159], Joon Ho Kang[76], Pamela K Keel[160], Jaeyoung Kim[161,162], Dongjun Kim[76], Tilo Kircher[163], George Kirov[94], Julia Kraft[32,33,164], John Kramer[165], Henry R Kranzler[166,167], Po-Hsiu Kuo[168], Siim Kurvits[169], Dongbing Lai[100], Marilyn T Lake[170,171], Mikael Landén[158,18], Séverine Lannoy[9], Matthew H Law[172,173,174], Byung-Chul Lee[76], Young Kee Lee[76], Kelli Lehto[169], Daniel F Levey[63], Cathryn M Lewis[80,81], Qingqin S Li[175,176], Calwing Liao[33,177], Penelope A Lind[66,173,174], Christine Lochner[178], Adriana Lori[179], Hermine HM Maes[180,8,9,181], Jayant Mahadevan[182], Mirko Manchia[65,183], Becky Mars[184,185,186], Nicholas G Martin[66], Lourdes Martorell[187,188,189,190], Andrew M McIntosh[191], Shelley F McMain[192,193], Andrew McQuillin[194], Sarah E Medland[66,195,196], Philip B Mitchell[142], Esther Molina[197,69,146], Eric T Monson[12,198], Mary S Mufford[199,200], Gerard Muntané[187,188,189,190,201], Richard Musil[202,203], Woojae Myung[156,204], Ana Iulia Neagu[205,95], Trine T Nielsen[92,93], Markus M Nöthen[90], Yaira Z Nunez[133,206], John I Nurnberger Jr[207,100,208], Satoshi Okazaki[20], Catherine M Olsen[172,209], Roel A Ophoff[210,211], Michael J Owen[94,212,213], Pedro M Pan[14], Sergi Papiol[214,215], Juan C Pascual[216,217], George P Patrinos[218,219,220,221,222], Joanna M Pawlak[223,224], Brenda WJH Penninx[225], Ana M Pérez-Gutiérrez[226], Nader Perroud[227,228], Roseann E Peterson[35,37], Claudia Pisanu[229], Giorgio Pistis[230], Bernice Porjesz[231,232], Danielle Posthuma[51,233], Abigail Powers[234], Martin

Preisig[230], Meera Purushottam[235], Andreas Reif[236,237], Eva Z Reininghaus[87], Miguel E Rentería[66,174,173], Stephan Ripke[32,33,164], Michael A Ripperger[45], Margarita Rivera[238,69,146], Emily K Roberts[239], Gloria Roberts[142], Linn Rødevand[116], Stefan Roepke[58,240], Diego L Rovaris[241], Giovanni A Salum[242,243], Alan R Sanders[244,245], Marcos L Santoro[13], Chelsea Sawyers[9,246,8], Stephen W Scherer[247,248,249], Claudia Schilling[250], Christian Schmahl[251], Peter R Schofield[142], Thomas G Schulze[252,253,254], Laura J Scott[255,256], Alessandro Serretti[257,258], Alexey Shadrin[15], Toshiyuki Shirai[20], Olav B Smeland[117,148], Jordan W Smoller[259,260], Marcus Sokolowski[261], Edmund J Sonuga-Barke[262], Alessio Squassina[229], Anna Starnawska[92], Nils Eiel Steen[117,148,263], Dan J Stein[264], Frederike Stein[91], Murray B Stein[22], Fabian Streit[265,266,267,268], Reeteka Sud[269], Patrick F Sullivan[270,271], Chikashi Terao[21,272,273], Claudio Toma[120,142,274], Leonardo Tondo[275,276], Gustavo Turecki[277,75], Rudolf Uher[23], Robert J Ursano[278], Sandra Van der Auwera[279], Marquis P Vawter[280], Alja Videtic Paska[281], Elisabet Vilella[187,188,189,190], John B Vincent[282], Biju Viswanath[283], Vladimir Vladimirov[284,285,286], Danuta E Wasserman[261,287], Thomas W Weickert[288], David C Whiteman[172,289], Virginia L Willour[290,291], Erik D Wiström[292], Stephanie H Witt[267,293,268], Hong-Hee Won[294], Robyn E Wootton[295,152,153,296], Clement C Zai[297,282,298,299], Jian Zhang[35], Lea Zillich[267,300], CVEDA and MGL*, Genoplan Research Team*, International Borderline Genomics Consortium*, Ole A Andreassen[116,117,292,148], Abraham A Palmer[22,301], Sandra Sanchez-Roige[22,301], J John Mann[19,302], Nathan A Kimbrel[56,44], Allison E Ashley-Koch[130,11], Douglas M Ruderfer[303,45,304], Anna R Docherty[12,305,306], Niamh Mullins[2,1,3,4]

[1]Department of Genetics and Genomic Sciences, Icahn School of Medicine at Mount Sinai, New York, NY, USA
[2]Department of Psychiatry, Icahn School of Medicine at Mount Sinai, New York, NY, USA
[3]Charles Bronfman Institute for Personalized Medicine, Icahn School of Medicine at Mount Sinai, New York, NY, USA
[4]Department of Artificial Intelligence and Human Health, Icahn School of Medicine at Mount Sinai, New York, NY, USA
[5]Department of Psychiatry, Washington University School of Medicine, St Louis, MO, USA
[6]AI for Health Institute, Washington University School of Medicine, St Louis, MO, USA
[7]Department of Cellular, Molecular, and Genetic Medicine, Virginia Commonwealth University, Richmond, VA, USA
[8]Virginia Institute for Psychiatric and Behavioral Genetics, Virginia Commonwealth University, Richmond, VA, USA
[9]Department of Psychiatry, Virginia Commonwealth University, Richmond, VA, USA
[10]Cooperative Studies Program Epidemiology Center-Durham, Durham Veterans Affairs Health Care System, Durham, NC, USA
[11]Duke Molecular Physiology Institute, Duke University Medical Center, Durham, NC, USA
[12]Department of Psychiatry, University of Utah, Salt Lake City, UT, USA
[13]Disciplina de Biologia Molecular, Universidade Federal de São Paulo, São Paulo, SP, Brazil
[14]Laboratory of Integrative Neuroscience, Department of Psychiatry, Universidade Federal de São Paulo, São Paulo, SP, Brazil
[15]Centre for Precision Psychiatry, Institute of Clinical Medicine, University of Oslo, Oslo, Norway
[16]Centre for Precision Psychiatry, Division of Mental Health and Addiction, Oslo University Hospital, Oslo, Norway
[17]Department of Medical Biochemistry and Biophysics, Karolinska Institutet, Stockholm, Sweden
[18]Department of Medical Epidemiology and Biostatistics, Karolinska Institutet, Stockholm, Sweden
[19]Department of Psychiatry, Columbia University, New York, NY, USA
[20]Department of Psychiatry, Kobe University Graduate School of Medicine, Kobe, Hyogo, Japan
[21]Laboratory for Statistical and Translational Genetics, RIKEN Center for Integrative Medical Sciences, Yokohama, Japan

[22]Department of Psychiatry, University of California San Diego, La Jolla, CA, USA
[23]Department of Psychiatry, Dalhousie University, Halifax, Nova Scotia, Canada
[24]NIMH, National Institute of Mental Health, Klecany, Czech Republic
[25]Institute of Environmental Medicine, Karolinska Institutet, Stockholm, Sweden
[26]Department of Psychiatry, Rutgers University, New Brunswick, NJ, USA
[27]Rutgers Addiction Research Center, Rutgers University, New Brunswick, NJ, USA
[28]Department of Neurology, Klinikum rechts der Isar, School of Medicine, Technical University of Munich, Munich, Germany
[29]Department of Psychiatry, Hospital Universitario La Paz, IdiPaz, School of Medicine, Universidad Autónoma de Madrid, CIBERSAM, Madrid, Spain
[30]Department of Clinical Psychology, University of Amsterdam, Amsterdam, the Netherlands
[31]Academic Center for Trauma and Personality, Academic Center for Trauma and Personality, Amsterdam, the Netherlands
[32]Department of Psychiatry and Psychotherapy, Campus Mitte, Charité – Universitätsmedizin Berlin, Berlin, Germany
[33]Stanley Center for Psychiatric Research, Broad Institute of MIT and Harvard, Cambridge, MA, USA
[34]WHO Collaborating Centre for Research and Training in Mental Health, Neurosciences and Substance Abuse, Department of Psychiatry, University of Ibadan, Ibadan, Nigeria
[35]Department of Psychiatry and Behavioral Sciences, SUNY Downstate Health Sciences University, Brooklyn, NY, USA
[36]Department of Community Health Sciences, SUNY Downstate Health Sciences University, Brooklyn, NY, USA
[37]Institute for Genomics in Health, SUNY Downstate Health Sciences University, Brooklyn, NY, USA
[38]Department of Genetics, Institute of Biosciences, Universidade Federal do Rio Grande do Sul, Porto Alegre, RS, Brazil
[39]ADHD Outpatient Program & Developmental Psychiatry Program, Hospital de Clínicas de Porto Alegre, Universidade Federal do Rio Grande do Sul, Porto Alegre, RS, Brazil
[40]Department of Psychiatry, University of Münster, Münster, Germany
[41]Department of Psychiatry, University of Melbourne, Melbourne, Australia
[42]Research Service, Mid-Atlantic Mental Illness Research Education and Clinical Center, Durham, NC, USA
[43]Research Service, Durham VA Health Care System, Durham, NC, USA
[44]Department of Psychiatry and Behavioral Sciences, Duke University School of Medicine, Durham, NC, USA
[45]Department of Biomedical Informatics, Vanderbilt University Medical Center, Nashville, TN, USA
[46]Department of Morphology and Genetics, Universidade Federal de São Paulo, São Paulo, SP, Brazil
[47]Institute of Epidemiology and Social Medicine, University of Münster, Münster, Germany
[48]Department of Quantitative Health Sciences, Mayo Clinic, Rochester, MN, USA
[49]Department of Psychiatry and Psychology, Mayo Clinic, Rochester, MN, USA
[50]Department of Psychological & Brain Sciences, Washington University in Saint Louis, St Louis, MO, USA
[51]Department of Complex Trait Genetics, Vrije Universiteit Amsterdam, Amsterdam, The Netherlands
[52]Amsterdam Reproduction & Development Research Institute, Vrije Universiteit Amsterdam, Amsterdam, The Netherlands
[53]Department of Biomedicine - Human Genetics, Aarhus University, Aarhus, Denmark
[54]The Lundbeck Foundation Initiative for Integrative Psychiatric Research, iPSYCH, Aarhus,

Aarhus, Denmark
[55]Center for Genomics and Personalized Medicine, Aarhus, Aarhus, Denmark
[56]VA Mid-Atlantic Mental Illness Research, Education, and Clinical Center, Durham VA Medical Center, Durham, NC, USA
[57]Department of Psychology, Georgetown University, Washington, D.C., USA
[58]Department of Psychiatry and Neuroscience, Charité - Universitätsmedizin Berlin, Berlin, Germany
[59]Department of Psychiatry, Federal University of São Paulo -UNIFESP, São Paulo, SP, Brasil
[60]Ame sua Mente, Instituto Ame sua Mente, São Paulo, SP, Brasil
[61]Department of Genes and Environment, Max Planck Institute of Psychiatry, Munich, Germany
[62]Institute of Psychiatric Phenomics and Genomics (IPPG), LMU University Hospital, LMU Munich, Munich, Germany
[63]Department of Psychiatry, Yale University, New Haven, CT, USA
[64]VA CT Healthcare System, West Haven, CT, USA
[65]Section of Psychiatry, Department of Medical Sciences and Public Health, University of Cagliari, Cagliari, Italy
[66]Brain and Mental Health Research Program, QIMR Berghofer Medical Research Institute, Brisbane, QLD, Australia
[67]Faculty of Health, Medicine and Behavioural Sciences, The University of Queensland, Brisbane, QLD, Australia
[68]Department of Psychiatry, University of Granada, Granada, Spain
[69]Institute of Neurosciences “Federico Olóriz”, Biomedical Research Center (CIBM), University of Granada, Granada, Spain
[70]Department of Mental Health, Clinico San Cecilio University Hospital, Granada, Spain
[71]Group 5, Instituto Biosanitario de Granada, Granada, Spain
[72]Institute of Epidemiology and Preventive Medicine, National Taiwan University, Taipei, Taiwan
[73]Université Paris Cité, NeuroDiderot (INSERM U1141), Institut Pasteur (CNRS UMR3571), Paris, France
[74]Hôpital Sainte-Anne, GHU Paris Psychiatrie et Neurosciences, Paris, France
[75]Department of Psychiatry, McGill University, Montreal, Quebec, Canada
[76]Genoplan, Genoplan RnD Division, Seoul, Republic of Korea
[77]Department of Biomedicine, University of Basel, Basel, Switzerland
[78]Institute of Medical Genetics and Pathology, University Hospital Basel, Basel, Switzerland
[79]Institute of Neuroscience and Medicine (INM-1), Research Center Juelich, Juelich, Germany
[80]Social, Genetic and Developmental Psychiatry Centre, King's College London, London, UK
[81]Institute of Psychiatry, Psychology & Neuroscience, King's College London, London, UK
[82]National Institute for Health Research Biomedical Research Centre, South London and Maudsley National Health Service Trust, London, UK
[83]Department of Neurobiology, University of Utah, Salt Lake City, UT, USA
[84]Department of Biomedical Informatics, University of Utah, Salt Lake City, UT, USA
[85]Department of Internal Medicine, University of Utah, Salt Lake City, UT, USA
[86]Department of Psychiatry, University of Vermont, Burlington, VT, USA
[87]Department of Psychiatry and Psychotherapeutic Medicine, Medical University of Graz, Graz, Austria
[88]Institute for Translational Psychiatry, University of Münster, Münster, Germany
[89]Medical School and University Medical Center OWL, Protestant Hospital of the Bethel Foundation, Department of Psychiatry, University of Bielefeld, Bielefeld, Germany
[90]Institute of Human Genetics, University of Bonn, School of Medicine & University Hospital Bonn, Bonn, Germany
[91]Department of Psychiatry and Psychotherapy, University of Marburg, Marburg, Germany
[92]Department of Biomedicine, Aarhus University, Aarhus, Denmark

[93]The Novo Nordisk Foundation Center for Genomic Mechanisms of Disease, Broad Institute of MIT and Harvard, Cambridge, MA, USA
[94]Division of Psychological Medicine and Clinical Neurosciences, Cardiff University, Cardiff, UK
[95]Department of Cellular and Molecular Pathology, Ştefan S. Nicolau Institute of Virology, Bucharest, Romania
[96]First Department of Psychiatry, Eginition Hospital, National and Kapodistrian University of Athens, Athens, Greece
[97]Department of Medical Genetics, Oslo University Hospital and University of Oslo, Oslo, Norway
[98]Centre for Precision Psychiatry, Division of Mental Health and Addiction, Oslo University Hospital and University of Oslo, Oslo, Norway
[99]Department of Biochemistry, Molecular Biology and Pharmacology, Indiana University School of Medicine, Indianapolis, IN, USA
[100]Department of Medical and Molecular Genetics, Indiana University School of Medicine, Indianapolis, IN, USA
[101]Danish Research Institute for Suicide Prevention, Mental Health Centre Copenhagen, Copenhagen, Denmark
[102]Copenhagen Research Centre for Mental Health, Mental Health Centre Copenhagen, Copenhagen, Denmark
[103]Centre of Mental Health Research, Australian National University, Canberra, ACT, Australia
[104]Department of Mental Health, Johns Hopkins Bloomberg School of Public Health, Baltimore, MD, USA
[105]Department of Consultation Psychiatry and Psychosomatics, University Hospital Zurich, Zurich, Switzerland
[106]Department of Psychiatry and Psychotherapy, LMU University Hospital, Munich, Germany
[107]Department of Psychiatry and Psychotherapy, Max Planck Institute of Psychiatry, Munich, Germany
[108]site Munich/Augsburg, German Center for Mental Health (DZPG), Germany
[109]Department of Medicine, University of Udine, Udine, Italy
[110]Department of Human Genetics, Radboud university medical center, Nijmegen, The Netherlands
[111]Department of Psychiatry, School of Medicine, National and Kapodistrian University of Athens, Athens, Greece
[112]Department of Clinical Psychology, University Hospital of Bellvitge-IDIBELL, Barcelona, Spain
[113]Clinical Sciences, School of Medicine and Health Sciences, University of Barcelona, Barcelona, Spain
[114]CIBER de Fisiopatología de la Obesidad y Nutrición (CIBERobn), Instituto de Salud Carlos III, Barcelona, Spain
[115]Institute of Neuroscience and Medicine (INM-1), Research Center Jülich, Jülich, Germany
[116]Centre for Precision Psychiatry, University of Oslo, Oslo, Norway
[117]Division of Mental Health and Addiction, Oslo University Hospital, Oslo, Norway
[118]Department of Pharmacy, Section for Pharmacology and Pharmaceutical Biosciences, University of Oslo, Oslo, Norway
[119]Department of Psychiatry and Behavioral Sciences, University of Texas Health Science Center at Houston, Houston, TX, USA
[120]Neuroscience Research Australia, Sydney, NSW, Australia
[121]School of Biomedical Sciences, Faculty of Medicine and Health, University of New South Wales, Sydney, NSW, Australia
[122]Pharmaceutical Sciences and Experimental Therapeutics, University of Iowa, Iowa City, IA, USA
[123]Department of Psychiatry, University of Iowa, Iowa City, IA, USA

[124]Iowa Neuroscience Institute, University of Iowa, Iowa City, IA, USA
[125]Department of Biostatistics, Columbia University, New York, NY, USA
[126]Veterans Affairs Connecticut Healthcare System, West Haven, CT, USA
[127]Environment and Health over the lifecourse, Barcelona Institute for Global Health (ISGlobal), Barcelona, Catalonia, Spain
[128]Department of Medicine and Life Sciences, Universitat Pompeu Fabra (UPF), Barcelona, Catalonia, Spain
[129]CIBER Epidemiología y Salud Pública (CIBERESP), Spain
[130]Department of Medicine, Duke University, Durham, NC, USA
[131]Department of Psychiatry, University Hospital Wuerzburg, Wuerzburg, Germany
[132]Department of Psychiatry Helios Hildburghausen, University Hospital Wuerzburg, Wuerzburg, Germany
[133]Department of Psychiatry, Yale School of Medicine, New Haven, CT, USA
[134]Department of Genetics, Yale University School of Medicine, West Haven, CT, USA
[135]Department of Neuroscience, Yale University School of Medicine, West Haven, CT, USA
[136]Wu Tsai Institute, Yale University School of Medicine, West Haven, CT, USA
[137]Department of Psychiatry, VA Connecticut Healthcare System, West Haven, CT, USA
[138]Department of Psychology and Mental Health, University of Worcester, Worcester, UK
[139]GHU Paris Psychiatrie et Neurosciences (Hôpital Sainte-Anne, CMME), Université Paris Cité, Paris, France
[140]Institute of Psychiatry and Neurosciences of Paris (IPNP), INSERM U1266, Paris, France
[141]Department of Psychiatry and Psychotherapy, University Medicine Greifswald, Greifswald, Mecklenburg - Vorpommern, Germany
[142]Discipline of Psychiatry and Mental Health, School of Clinical Medicine, Faculty of Medicine and Health, University of New South Wales, Sydney, NSW, Australia
[143]Psychiatric Genetics Research Unit, Alexandru Obregia Clinical Psychiatric Hospital, Bucharest, Romania
[144]Division of Human Genetics, Department of Psychiatry, Yale University School of Medicine, New Haven, CT, USA
[145]Department of Psychiatry, Veterans Affairs Connecticut Healthcare Center, West Haven, CT, USA
[146]Instituto de Investigación Biosanitaria, Ibs Granada, Granada, Spain
[147]Unidad de Gestión Clínica de Salud Mental, Hospital Regional Universitario de Málaga, Instituto de Investigación Biomédica de Málaga y Plataforma en Nanomedicina - IBIMA Plataforma Bionand, Málaga, Spain
[148]Institute of Clinical Medicine, University of Oslo, Oslo, Norway
[149]Department of General Psychiatry, Heidelberg University, Heidelberg, Germany
[150]Department of Psychiatry, Univ of CT School of Medicine, Farmington, CT, USA
[151]Integrative Medicine, National Institute of Mental Health and Neuro Sciences, Bengaluru, India
[152]Research Department, Lovisenberg Diaconal Hospital, Oslo, Norway
[153]PsychGen Center for Genetic Epidemiology and Mental Health, Norwegian Institute of Public Health, Oslo, Norway
[154]Department of Psychology, University of Oslo, Oslo, Norway
[155]Department of Health Science and Technology, Seoul National University, Seoul, Republic of Korea
[156]Department of Neuropsychiatry, Seoul National University Bundang Hospital, Seongnam, Republic of Korea
[157]Three Counties Medical School, University of Worcester, Worcester, UK
[158]Institute of Neuroscience and Physiology, University of Gothenburg, Gothenburg, Sweden
[159]Department of Anesthesiology, Mass General Brigham, Boston, MA, USA

[160]Department of Psychology, Florida State University, Tallahassee, FL, USA
[161]Department of Digital Health, Samsung Advanced Institute for Health Sciences and Technology (SAIHST), Sungkyunkwan University, Samsung Medical Center, Seoul, South Korea, Seoul, Republic of Korea
[162]Department of Neuropsychiatry, Seoul National University Bundang Hospital, Seongnam, South Korea, Seongnam, Republic of Korea
[163]Department of Psychiatry, Marburg University, Marburg, Germany
[164]partner site Berlin/Potsdam, German Center for Mental Health (DZPG), Germany
[165]Department of Psychiatry, University of Iowa Carver College of Medicine, Iowa City, IA, USA
[166]Department of Psychiatry, University of Pennsylvania Perelman School of Medicine, Philadelphia, PA, USA
[167]Mental Illness Research, Education and Clinical Center, Crescenz Veterans Affairs Medical Center, Philadelphia, PA, USA
[168]Department of Public Health & Institute of Epidemiology and Preventive Medicine, National Taiwan University, Taipei, Taiwan
[169]Estonian Genome Centre, Institute of Genomics, University of Tartu, Tartu, Estonia
[170]Department of Psychiatry and Neuroscience Institute, Faculty of Health Sciences, University of Cape Town, Cape Town, South Africa
[171]Department of Paediatrics and Child Health, South African Medical Research Council (SAMRC) Unit on Child and Adolescent Health, University of Cape Town, Cape Town, South Africa
[172]Population Health Research Program, QIMR Berghofer Medical Research Institute, Brisbane, QLD, Australia
[173]School of Biomedical Sciences, Faculty of Health, Queensland University of Technology, Brisbane, QLD, Australia
[174]School of Biomedical Sciences, Faculty of Health, Medicine and Behavioural Sciences, The University of Queensland, Brisbane, QLD, Australia
[175]Department of Neuroscience, Janssen Research & Development, LLC, Titusville, NJ, USA
[176]Preclinical Department, CHDI Management, Inc., Princeton, NJ, USA
[177]Analytical and Translational Genetics Unit, Massachusetts General Hospital, Cambridge, MA, USA
[178]SAMRC Unit on Risk & Resilience in Mental Disorders, Department of Psychiatry, Stellenbosch University, Cape Town, South Africa
[179]Department of Population Science, American Cancer Society, Atlanta, GA, USA
[180]Department of Cellular, Molecular and Genetic Medicine, Virginia Commonwealth University, Richmond, VA, USA
[181]Massey Cancer Center, Virginia Commonwealth University, Richmond, VA, USA
[182]Department of Psychiatry, National Institute of Mental Health and Neurosciences, Bangalore, India
[183]Department of Pharmacology, Dalhousie University, Halifax, Canada
[184]Centre of Academic Mental Health, University of Bristol, Bristol, UK
[185]Department of Population Health Sciences, University of Bristol, Bristol, UK
[186]National Institute for Health Research Biomedical Research Centre, University Hospitals Bristol and Weston NHS Foundation Trust and University of Bristol, Bristol, UK
[187]Research Area, Hospital Universitari Institut Pere Mata, Reus, Spain
[188]Genetics and Environment in Psychiatry Research Group, Institut d'Investigació Sanitària Pere Virgili-CERCA, Reus, Spain
[189]Medicine and Surgery Department, Faculty of Medicine and Health Sciences, Universitat Rovira i Virgili (URV), Reus, Spain
[190]Centro de Investigación Biomédica en Red de Salud Mental (CIBERSAM), Instituto de Salud Carlos III, Madrid, Spain

[191]Institute for Cardiovascular and Neuroscience Research, University of Edinburgh, Edinburgh, UK
[192]BPD Clinic, Centre for Addiction and Mental Health, Toronto, Ontario, Canada
[193]Department of Psychiatry, University of Toronto, Toronto, Ontario, Canada
[194]Division of Psychiatry, University College London, London, UK
[195]School of Psychology, The University of Queensland, Brisbane, QLD, Australia
[196]School of Psychology and Counselling, Queensland University of Technology, Brisbane, QLD, Australia
[197]Department of Nursing, Faculty of Health Sciences, University of Granada, Granada, Spain
[198]Huntsman Mental Health Institute, University of Utah, Salt Lake City, UT, USA
[199]Division of Human Genetics, Department of Pathology, University of Witwatersrand, Johannesburg, Gauteng, South Africa
[200]Department of Psychiatry and Mental Health, University of Cape Town, Cape Town, Western Cape, South Africa
[201]Institut de Biologia Evolutiva (IBE, UPF-CSIC), Department of Medicine and Life Sciences, Universitat Pompeu Fabra, Parc de Recerca Biomèdica de Barcelona (PRBB), Barcelona, Spain
[202]Psychiatric Clinic, Ludwig Maximilian University, Munich, Germany
[203]Psychosomatic Clinic, Oberberg Fachklinik Bad Tölz, Bad Tölz, Germany
[204]Department of Neuropsychiatry, Seoul National University College of Medicine, Seoul, Republic of Korea
[205]Department of Virology, Carol Davila University of Medicine and Pharmacy, Bucharest, Romania
[206]Department of Research, West Haven VA Medical Center, West Haven, CT, USA
[207]Department of Psychiatry, Indiana University School of Medicine, Indianapolis, IN, USA
[208]Stark Neurosciences Research Institute, Indiana University School of Medicine, Indianapolis, IN, USA
[209]Frazer Institute, The University of Queensland, Brisbane, QLD, Australia
[210]Center for Neurobehavioral Genetics, Semel Institute for Neuroscience and Human Behavior, University of California Los Angeles, Los Angeles, CA, USA
[211]Department of Human Genetics, University of California Los Angeles, Los Angeles, CA, USA
[212]Centre for Neuropsychiatric Genetics and Genomics, Cardiff University, Cardiff, UK
[213]Neurosciences and Mental Health Innovation Institute, Cardiff University, Cardiff, UK
[214]Institute of Psychiatric Phenomics and Genomics (IPPG), LMU University Hospital, Munich, Germany
[215]Department of Clinical Translation, Max Planck Institute of Psychiatry, Munich, Germany
[216]Mental Health and Psychiatry Department, Vic Hospital Consortium, Vic, Barcelona, Spain
[217]Instituto de Salud Carlos III, Centro de Investigación Biomédica en Red de Salud Mental, Instituto de Salud Carlos III, Madrid, Spain
[218]Department of Pharmacy, Patras, Greece, University of Patras School of Health Sciences, Patras, Greece
[219]Department of Genetics and Genomics, United Arab Emirates University College of Medicine and Health Sciences, Al Ain, Abu Dhabi, UAE
[220]Zayed Center for Health Sciences, United Arab Emirates University, Al Ain, Abu Dhabi, UAE
[221]Department of Pathology, Clinical Bioinformatics Unit, Erasmus University Medical Center, Faculty of Medicine and Health Sciences, Rotterdam, The Netherlands
[222]Hellenic Pasteur Institute, Athens, Greece
[223]Department of Psychiatric Genetics, Poznan University of Medical Sciences, Poznan, Poland
[224]Department of Psychiatry, Poznan University of Medical Sciences, Poznan, Poland
[225]Department of Psychiatry, Amsterdam UMC, Vrije Universiteit, Amsterdam, The Netherlands
[226]Department of Biochemistry and Molecular Biology II, University of Granada, Granada, Spain

[227]Department of Psychiatry, University of Geneva, Faculty of Medicine, Geneva, Switzerland
[228]Medical Direction, Geneva University Hospitals, Division of Institutional Measures, Geneva, Switzerland
[229]Department of Biomedical Sciences, University of Cagliari, Cagliari, Italy
[230]Department of Psychiatry, Lausanne University Hospital and University of Lausanne, Lausanne, Vaud, Switzerland
[231]Department of Psychiatry, SUNY Downstate Health Sciences University, Brooklyn, NY, USA
[232]Henri Begleiter Neurodynamics Lab, SUNY Downstate Health Sciences University, Brooklyn, NY, USA
[233]Department of Child and Adolescent Psychiatry, Amsterdam University Medical Center, Amsterdam, The Netherlands
[234]Department of Psychiatry and Behavioral Sciences, Emory University, Atlanta, GA, USA
[235]Molecular Genetics Laboratory, Department of Psychiatry, National Institute of Mental Health and Neurosciences, Bengaluru, Karnataka, India
[236]Department of Psychiatry, Psychosomatic Medicine and Psychotherapy, University Hospital Frankfurt – Goethe University, Frankfurt am Main, Germany, Frankfurt am Main, Hesse, Germany
[237]Fraunhofer Institute for Translational Medicine and Pharmacology ITMP, Theodor-Stern-Kai 7, 60596 Frankfurt am Main, Germany, Frankfurt am Main, Hesse, Germany
[238]Department of Biochemistry and Molecular Biology II, Faculty of Pharmacy, University of Granada, Granada, Spain
[239]Department of Biostatistics, University of Iowa, Iowa City, IA, USA
[240]Department of Psychiatry, Psychosomatic Medicine and Psychotherapy, Oberbergkliniken, Berlin Brandenburg, Germany
[241]Laboratory of Physiological Genomics of Neurodevelopment (PhysioGen Lab), Instituto de Ciencias Biomedicas, Universidade de São Paulo, São Paulo, SP, Brazil
[242]Department of Psychiatry, Universidade Federal do Rio Grande do Sul, Porto Alegre, RS, Brazil
[243]Global Programs, Child Mind Institute, New York, USA
[244]Department of Psychiatry and Behavioral Sciences, Endeavor Health, Evanston, IL, USA
[245]Department of Psychiatry and Behavioral Neuroscience, University of Chicago, Chicago, IL, USA
[246]Division of Child and Adolescent Psychiatry, Virginia Commonwealth University, Richmond, VA, USA
[247]Genetics and Genome Biology Program, The Hospital for Sick Children, Toronto, Ontario, Canada
[248]The Centre for Applied Genomics (TCAG), The Hospital for Sick Children, Toronto, Ontario, Canada
[249]Department of Molecular Genetics and The McLaughlin Centre, University of Toronto, Toronto, Ontario, Canada
[250]Department of Psychiatry and Psychotherapy, Sleep Laboratory, Medical Faculty Mannheim, University of Heidelberg, Central Institute of Mental Health, Mannheim, Germany
[251]Department of Psychosomatic Medicine and Psychotherapy, Central Institute of Mental Health, Mannheim, Germany
[252]Institute of Psychiatric Phenomics and Genomics (IPPG), University Hospital, LMU Munich, Munich, Bavaria, Germany
[253]Department of Psychiatry and Behavioral Sciences, Norton College of Medicine, SUNY Upstate Medical University, Syracuse, NY, USA
[254]Department of Psychiatry and Behavioral Sciences, The Johns Hopkins University, Baltimore, MD, USA
[255]Department of Biostatistics, School of Public Health, University of Michigan, Ann Arbor, MI,

USA
[256]Center for Statistical Genetics, University of Michigan, Ann Arbor, MI, USA
[257]Department of Medicine and Surgery, Kore University of Enna, Enna, Italy
[258]Oasi Research Institute-IRCCS, Troina, italy
[259]Center for Precision Psychiatry, Massachusetts General Hospital, Boston, MA, USA
[260]Psychiatric and Neurodevelopmental Genetics Unit, Center for Genomic Medicine, Massachusetts General Hospital, Boston, MA, USA
[261]National Centre for Suicide Research and Prevention of Mental Ill-Health (NASP), LIME, Karolinska Institute, Stockholm, Sweden
[262]Department of Child and Adolescent Psychiatry, King's College London, London, UK
[263]Division of Mental Health and Substance Abuse, Diakonhjemmet Hospital, Oslo, Norway
[264]SAMRC Unit on Risk & Resilience in Mental Disorders, Department of Psychiatry & Neuroscience Institute, University of Cape Town, Cape Town, South Africa
[265]Hector Institute for Artificial Intelligence in Psychiatry, Central Institute of Mental Health, Medical Faculty Mannheim, Heidelberg University, Mannheim, Germany
[266]Department of Psychiatry and Psychotherapy, Central Institute of Mental Health, Medical Faculty Mannheim, Heidelberg University, Mannheim, Germany
[267]Department of Genetic Epidemiology in Psychiatry, Central Institute of Mental Health, Medical Faculty Mannheim, Heidelberg University, Mannheim, Germany
[268]German Center for Mental Health (DZPG), Partner Site Mannheim - Heidelberg - Ulm, Germany
[269]Center for Brain and Mind, Department of Psychiatry, National Institute of Mental Health and Neurosciences (NIMHANS), Bangalore, India
[270]Department of Genetics, University of North Carolina, Chapel Hill, NC, USA
[271]Department of Psychiatry, University of North Carolina, Chapel Hill, NC, USA
[272]Clinical Research Center, Shizuoka General Hospital, Shizuoka, Japan
[273]Applied Genetics, The School of Pharmaceutical Sciences, University of Shizuoka, Shizuoka, Japan
[274]Department of Molecular Neuropathology, Centro de Biología Molecular Severo Ochoa, Consejo Superior de Investigaciones Científicas (CSIC) - Universidad Autónoma de Madrid (UAM), Madrid, Spain
[275]Department of Psychiatry, McLean Hospital - Harvard Medical School, Boston, MA, USA
[276]Department of Psychiatry, Mood Disorders Centro Lucio Bini, Rome, Italy
[277]Douglas Institute, McGill University, Montreal, Quebec, Canada
[278]Center for the Study of Traumatic Stress (CSTS), Department of Psychiatry, Uniformed Services University of the Health Sciences (USUHS), Bethesda, MD, USA
[279]Department of Psychiatry, University Medicine Greifswald, Greifswald, Germany
[280]Department of Psychiatry and Human Behavior, University of California, Irvine, Irvine, CA, USA
[281]Faculty of Medicine, Institute of Biochemistry and Molecular Genetics, University of Ljubljana, Ljubljana, Slovenia
[282]Molecular Brain Science, Centre for Addiction and Mental Health, Toronto, Ontario, Canada
[283]Department of Psychiatry, National Institute of Mental Health and Neurosciences, Bangalore, Karnataka, India
[284]Department of Psychiatry, The University of Arizona, Phoenix, AZ, USA
[285]GDIP, The University of Arizona, Phoenix, AZ, USA
[286]The Lieber Institute for Brain Development, The Lieber Institute for Brain Development, Baltimore, MD, USA
[287]NASP, Centrum för Hälsoekonomi,Informatik och Vårdforskning (CHIS), Stockholms läns sjukvårdsområde SLSO, Region Stockholm, Sweden
[288]Department of Psychiatry, SUNY Upstate Medical University, Syracuse, NY, USA

[289]School of Public Health, The University of Queensland, Brisbane, QLD, Australia
[290]Department of Psychiatry and Pediatrics, University of Iowa, Iowa City, IA, USA
[291]Iowa City Health Care System, Department of Veterans Affairs, Iowa City, IA, USA
[292]Centre for Precision Psychiatry, Oslo University Hospital, Oslo, Norway
[293]Biobank of the Center for Innovative Psychiatric and Psychotherapeutic Research, Central Institute of Mental Health, Medical Faculty Mannheim, Heidelberg University, Mannheim, Germany
[294]Samsung Advanced Institute for Health Sciences and Technology (SAIHST), Sungkyunkwan University, Seoul, Republic of Korea
[295]School of Psychological Science, University of Bristol, Bristol, UK
[296]Integrative Epidemiology Unit, University of Bristol, Bristol, UK
[297]Tanenbaum Centre for Pharmacogenetics, Centre for Addiction and Mental Health, Toronto, Ontario, Canada
[298]Campbell Family Mental Health Research Institute, Centre for Addiction and Mental Health, Toronto, Ontario, Canada
[299]Department of Psychiatry, Institute of Medical Science, Laboratory Medicine and Pathobiology; Dalla Lana School of Public Health, University of Toronto, Toronto, Ontario, Canada
[300]Department of Psychiatry and Psychotherapy, University Medical Center Freiburg, Freiburg, Germany
[301]Institute for Genomic Medicine, University of California San Diego, La Jolla, CA, USA
[302]Department of Radiology, Columbia University, New York, NY, USA
[303]Center for Digital Genomic Medicine, Division of Genetic Medicine, Department of Medicine, Vanderbilt University Medical Center, Nashville, TN, USA
[304]Department of Psychiatry and Behavioral Sciences, Vanderbilt University Medical Center, Nashville, TN, USA
[305]University of Utah Health Sciences, Clinical and Translational Science Institute, Salt Lake City, UT, USA
[306]Suicide Research and Prevention, Huntsman Mental Health Institute, Salt Lake City, UT, USA
